# Graph Theoretical Measures of Fast Ripples Support the Epileptic Network Hypothesis

**DOI:** 10.1101/2022.01.20.22269492

**Authors:** Shennan A Weiss, Tomas Pastore, Iren Orosz, Daniel Rubinstein, Richard Gorniak, Zachary Waldman, Itzhak Fried, Chengyuan Wu, Ashwini Sharan, Diego Slezak, Gregory Worrell, Jerome Engel, Michael R. Sperling, Richard J Staba

## Abstract

The epileptic network hypothesis and epileptogenic zone (EZ) hypothesis are two theories of ictogenesis. The network hypothesis posits that coordinated activity among interconnected nodes produces seizures. The EZ hypothesis posits that distinct regions are necessary and sufficient for seizure generation. High-frequency oscillations (HFOs), and particularly fast ripples (FR), are thought to be biomarkers of the EZ. We sought to test these theories by comparing HFO rates and networks in surgical responders and non-responders, with no appreciable change in seizure frequency or severity, within a retrospective cohort of 48 patients implanted with stereo-EEG electrodes. We recorded inter-ictal activity during non-rapid eye movement sleep and semi-automatically detected and quantified HFOs. Each electrode contact was localized in normalized coordinates. We found that the accuracy of seizure-onset zone (SOZ) electrode contact classification using HFO rates (ripples 80-250 Hz, and FR 250-600 Hz) was not significantly different in surgical responders and non-responders, suggesting that in non-responders the EZ partially encompassed the SOZ(s) (p>0.05). We also found that in the responders, FR on oscillations exhibited a higher spectral content in the SOZ compared to the non-SOZ (p<1e-5). In contrast, in the non-responders the FR had a lower spectral content in the SOZ (p<1e-5). We constructed two different networks of FR with a spectral content > 350 Hz. The first was a rate-distance network that multiplied the Euclidian distance between FR generating contacts by the average rate of FR in the two contacts. The radius of the rate-distance network, which excluded SOZ nodes, discriminated non-responders, including patients not offered resection or responsive neurostimulation (RNS) due to diffuse multi-focal onsets, with an accuracy of 0.77 [95% confidence interval (CI) 0.56-0.98]. The second FR network was constructed using the mutual information (MI) between the timing of the events to measure functional connectivity. For most non-responders, this network had a longer characteristic path length, lower mean local efficiency in the non-SOZ, and a higher nodal strength among non-SOZ nodes relative to SOZ nodes. The graphical theoretical measures from the rate-distance and MI networks of 22 non-RNS treated patients was used to train a support vector machine, which when tested on 13 distinct patients classified non-responders with an accuracy of 0.92 [95% CI 0.75-1]. These results support the epileptic network hypothesis, refute the EZ hypothesis, and suggest that surgical non-responders, and patients not selected for resection or RNS, exhibit a decentralized FR network consisting of widely distributed, hyperexcitable FR generating nodes.

**Abbreviated Summary:** Weiss et al., report that fast ripples form a correlational network. In poor epilepsy surgery candidates, and patients that don’t respond to surgery, the networks are widespread, highly active, and decentralized. This can be used to plan effective surgeries and supports the theory that seizures require a coordinated epileptic network.

Graphical abstract

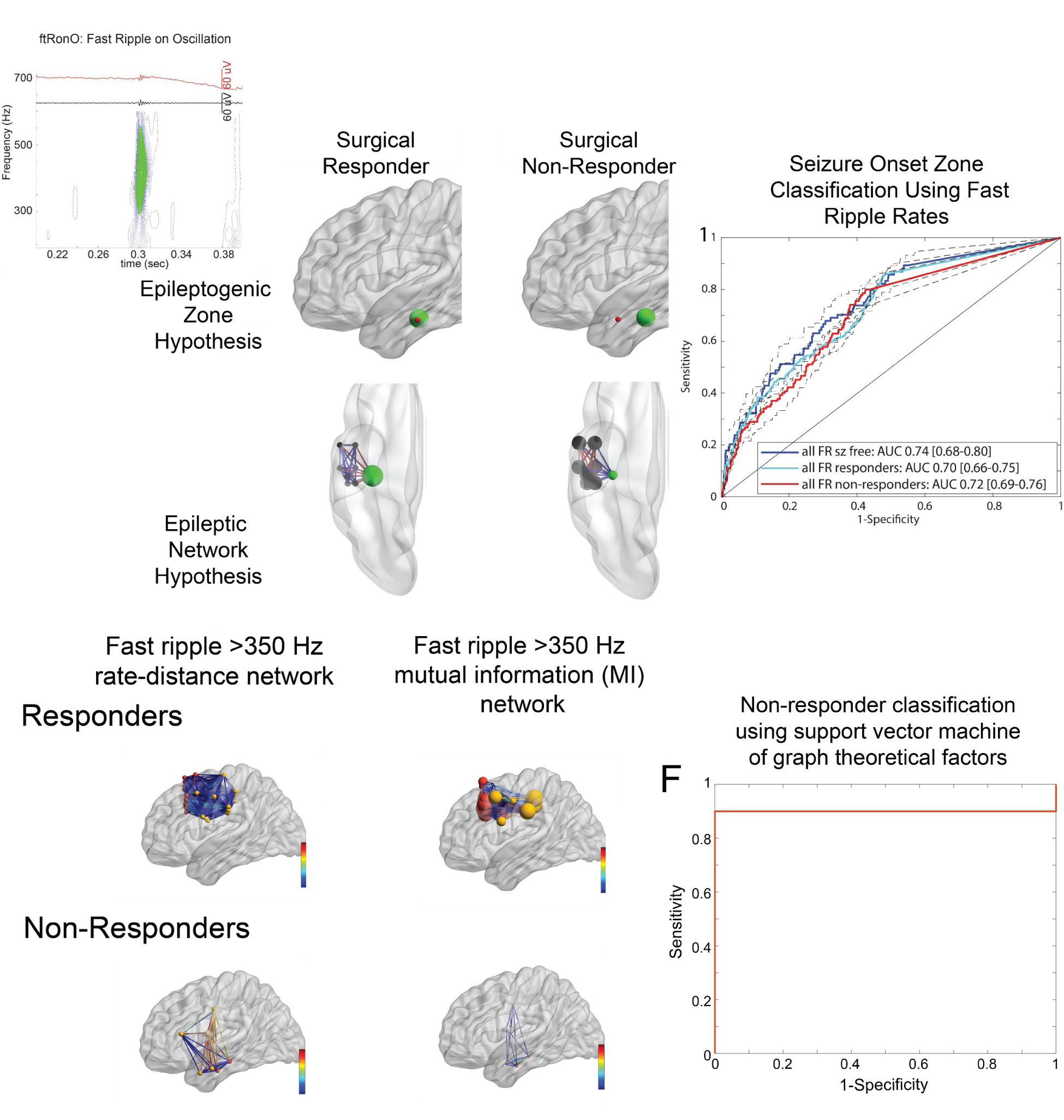

## Introduction

Different theoretical frameworks of ictogenesis have been constructed to understand the mechanisms generating seizures and interpret seizure outcome after therapy to control or eliminate seizures. Two predominant theories of ictogenesis are the epileptogenic zone hypothesis^1–3^ and the epileptic network hypothesis^4–10^. The epileptogenic zone is defined as “the region that is indispensable for the generation of seizures, or the area of cortex that is necessary and sufficient for initiating seizures and whose removal (or disconnection) is necessary for complete abolition of seizures”^1^. The EZ is inferred retrospectively as the region within the resection margins in patients with seizure free outcomes^1, 3^. The EZ, for example, might encompass the site of seizure onset (seizure onset zone or SOZ) or could reside within the SOZ as a focal MRI lesion, but in either case, the EZ must also be resected to achieve seizure freedom^2^ (Figure 1). Several EZs may exist independently, and in this case, patients are thought to have a worse outcome from epilepsy surgery^11^. Also, the EZ may be dynamic and new EZs could potentially develop after a surgery that targeted the initial EZ^12–14^.

**Figure 1:**
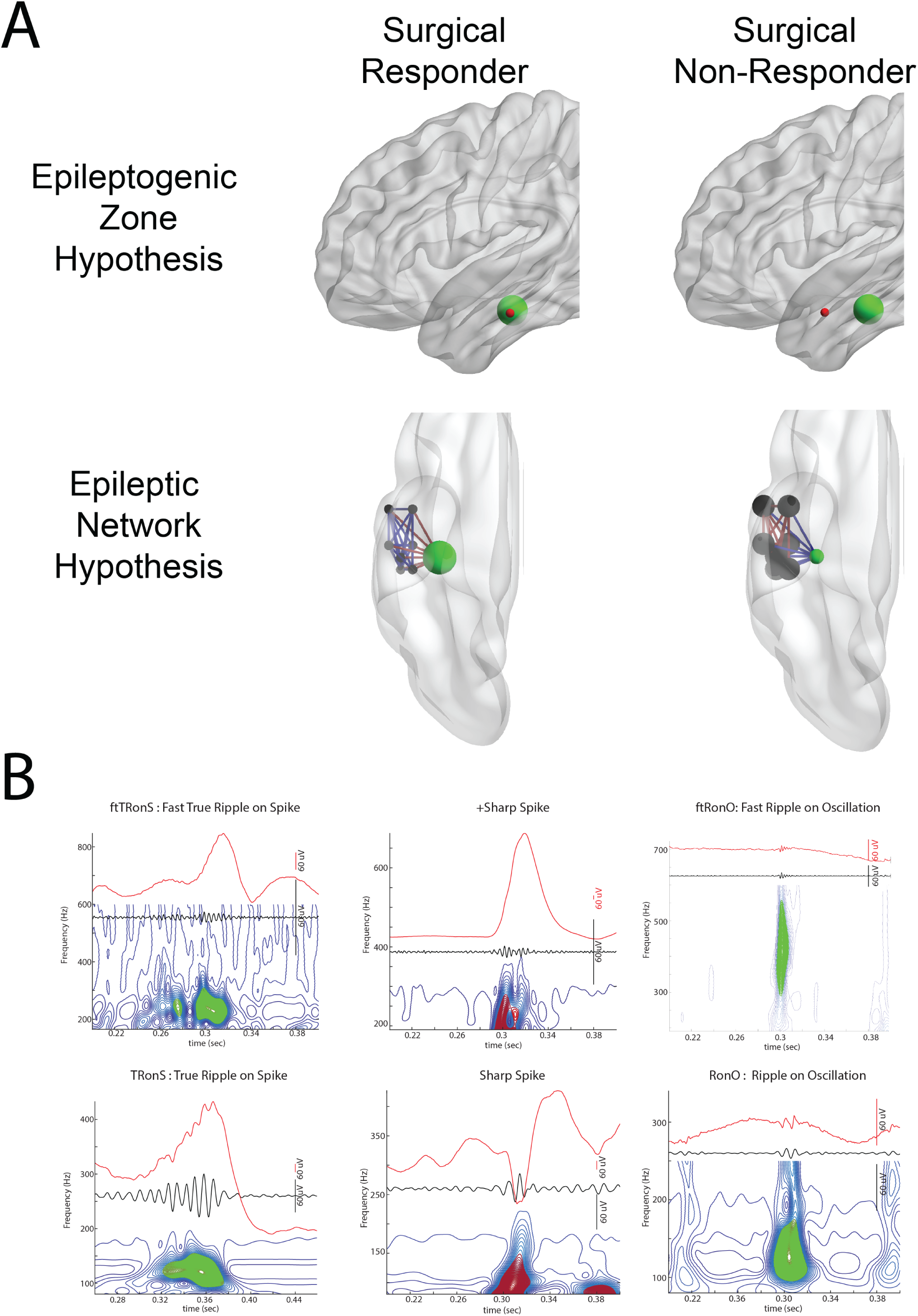
Illustration of two potential mechanisms accounting for epilepsy surgery failure (A) and HFO and spike biomarkers (B). (A) In the epileptogenic zone (EZ) hypothesis the EZ (red) is necessary and sufficient for seizure generation. When the EZ overlaps with the seizure onset zone (SOZ, green), and the SOZ is resected, the patient is a surgical responder. However, if the EZ is discordant with the SOZ, and the SOZ is resected instead of the EZ, the patient will be a surgical non-responder. In the epileptic network hypothesis, the nodes of the epileptic network (black) are connected to each other by weighted connections. If the SOZ node (green) is the most strongly connected (red edges) to the other nodes, then resecting the SOZ node alone will result in a surgical response. In contrast, if the non-SOZ nodes are most strongly connected with each other (red edges), and weakly connected (blue edges) with the SOZ node, the patient will be a surgical failure if only the SOZ node is resected. (B) Illustration of the HFO subtypes detected using the topographical analysis method. Each panel includes the iEEG trace (above), the 80-600 Hz band-pass filtered iEEG (middle), and the corresponding contour lines of isopower in the time-frequency spectrogram (below). Each contour line is shown in blue, groups of closed loop contours are in green, open loop contours are in dark red. Note that sharp spikes produce HFOs after band-pass filtering but no closed loop contours.

High-frequency oscillations (ripples: 80-250 Hz/fast ripple FR:250-600 Hz) are considered biomarkers of the EZ^15, 16^. FRs are closely linked to epileptogenesis^17, 18^ and in patients, specific to the SOZ and EZ and rare outside the EZ^19^. By contrast, ripples can be pathologic events^20^, but are important physiologic events in the normal hippocampus^21^ and other brain areas^19, 22^ In patient studies that investigate seizure outcome following epilepsy surgery, the resection of FR predicts seizure freedom better than ripples^23–29^. If regions generating high rates of FR are not resected, or identified^30^, seizure freedom is much less likely to be achieved^23–30^.

However, at the individual patient level, resection of FR sites does not always predict outcome^30–32^. Furthermore, in some seizure-free patients sites with high rates of FR do remain^30–32^.

Recently, it has also been suggested that HFOs sites are functionally connected which may be important in predicting response to epilepsy surgery^33–35^.

In contrast to the EZ hypothesis, the epileptic network hypothesis postulates that epileptogenesis and ictogenesis might be distributed and connected by functional and structural brain networks outside the SOZ. The initial area of apparent seizure involvement is not really an onset area, because “onset” could be expressed in any part of the network and might even vary from seizure to seizure in each patient. (Figure 1)^4, 36^. The pragmatic implications of the epileptic network hypothesis have been important in understanding and predicting the response to focal resection^37–39^, and responsive neurostimulation (RNS)^3, 40–42^. In contrast to resective surgery, RNS electrically modulates part(s) of the network and exhibits clinical benefits that slowly improve over time^40–42^. The epileptic network hypothesis has been investigated using graph theory, a mathematical method for describing both the global and local properties of networks consisting of nodes connected by edges^43^.

Structural and functional MRI studies as well as neurophysiologic studies using diverse approaches have provided strong evidence for the epileptic network hypothesis. Structural MRI studies have shown that patients with focal epilepsy have significantly lower fractional anisotropy in most fiber tracts, even those far from the presumed EZ^44^. Also distant from the EZ cortical thickness can be decreased^45^ and changes in a structural graph theoretical measure called node abnormality occur^46^. Graph theoretical measures of the networks characterized by resting state fMRI^5, 7, 9^, EEG^5, 7, 9, 37, 38, 47–52^, and MEG^53^ connectivity are altered in patients with focal epilepsy, both in the presumed EZ^37, 38, 47, 48, 50, 51^ and at distant sites^49, 52–54^. Ictal EEG functional connectivity networks have been constructed using the epileptogenicity index^8, 55–57^, and the coherence of the broad band EEG signal^39, 58, 59^. Studies using the epileptogenicity index have shown that a greater number of interconnected epileptogenic regions correlate with lower likelihood for seizure-free outcome^8, 55–57^, while studies using broad band coherence have shown that stronger opposing interactions between brain areas that lowers network synchrony also constrains seizure spread^59^. Furthermore, decreased synchronization at the time of seizure onset is predictive of good outcome^39^.

In this study, we evaluated the EZ and epileptic network hypotheses using inter-ictal HFOs recorded during non-REM sleep in surgical patients who responded to therapy, i.e., had reduction in seizures after treatment, and patients who had no change in seizure frequency or severity after treatment (non-responders). We postulated if the EZ hypothesis is correct and FR are a biomarker of the EZ, then classification accuracy of the SOZ using FR should be lower in non-responders than responders due to discordance between the location of SOZ and EZ. However, if the epileptic network hypothesis is correct, then the spatial distribution of FR-generating sites and connectivity between these sites, especially outside the SOZ, should be different between non-responders and responders. We used multi-site depth electrode recordings to construct FR networks, graph theory to characterize the network, and support vector machine (SVM) learning to predict response to surgery.

## Materials and methods

### 1. Patients

This retrospective study of diagnostic accuracy utilizing machine learning used consecutive recordings selected from 19 patients who underwent intracranial monitoring with depth electrodes between 2014-2018 at University of California Los Angeles (UCLA) and from 29 patients at Thomas Jefferson University (TJU) in 2016–2018 for the purpose of localization of the seizure onset zone. These patients were assigned to the training and testing sets of this study on the bases of data availability at the time of analysis and were not randomized. Patients had pre-surgical magnetic resonance imaging (MRI) for MRI-guided stereotactic electrode implantation, as well as a post-implant CT scan to localize the electrodes. All patients provided verbal and written consent prior to participating in this research, which was approved by the UCLA and TJU institutional review boards. The inclusion criteria were at least one night of intracranial recording at a 2000 Hz sampling rate uninterrupted by seizures and at least 4 h of interictal non-REM iEEG recordings. One to two days after implantation, for each patient a 10 to 60 minutes iEEG recording from all the depth electrodes that contained large amplitude, delta-frequency slow waves (i.e., non-REM sleep) was selected for analysis. Only iEEG that was free of low levels of muscle contamination and other artifacts was selected. Among all patients enrolled in the research study, recordings that met the inclusion criteria were available for approximately 60% of the UCLA patients, and 78% of the TJU patients. At UCLA research recordings were not always performed. No other patients were excluded on any other basis. The attending neurologist determined the seizure onset zone from visual inspection of video-EEG during the patient’s habitual seizures. The seizure onset zone was aggregated across all these seizures during the entire iEEG evaluation for each patient and did not include areas of early propagation. The non-SOZ included all remaining contacts and was often separated from the SOZ by sub-centimeter distances.

### 2. Electrode Localization

T1- and T2-weighted MRIs were obtained for each patient, prior to electrode implantation. FreeSurfer (http://surfer.nmr.mgh.harvard.edu/) was used on the T1-weighted MRI to construct individual subject brain surfaces and cortical parcellations according to the Desikan–Killiany atlas^60^. With the assistance of a neuroradiologist the Advanced Neuroimaging Tools ^61^ was used to coregister the post-implantation CT with the pre-implantation MRI, and the position of each electrode contact was localized to the Desikan-Killiany atlas. Then an in-house pipeline (https://github.com/pennmem/neurorad_pipeline) was used to transform the position of each electrode contact from individual subject space to an averaged FreeSurfer space with normalized Montreal Neurological Institute (MNI) coordinates (defined by the fsaverage brain).

### 3. EEG Recordings and HFO Detection

For each patient, clinical iEEG (0.1–600 Hz; 2000 samples per second) was recorded from 8-16 depth electrodes, each with 7 to 15 contacts, using a Nihon-Kohden 256-channel JE-120 long-term monitoring system (Nihon-Kohden America, Foothill Ranch, CA, U.S.A.). A larger number of electrodes with more contacts were implanted at TJU. The reference signal used for the recordings performed at UCLA was a scalp electrode position at Fz in the International 10-20 System. The reference signal used for the TJU recordings was an electrode in the white matter.

HFOs and sharp-spikes were detected in the non-REM sleep iEEG using previously published methods (https://gitlab.liaa.dc.uba.ar/)^29,62–65^ implemented in Matlab (Mathworks, Natick, MA, USA). In brief, the HFO detector reduced muscle and electrode artifacts in the iEEG recordings using an independent component analysis (ICA)-based algorithm^65^. Events were then detected, quantified, and classified in the referential and bipolar montage iEEG recordings per contact by utilizing a Hilbert detector followed by a topographic analysis of each event^29, 62, 65^ (Supplemental Methods). Following automatic detection of HFO and sharp-spikes, false detections of clear muscle and mechanical artifact were deleted by visual review in Micromed Brainquick (Venice, Italy).

### 4. Statistics, Graph Theoretical Measures, and Support Vector Machine (SVM)

Receiver operating characteristic curves were generated using the perfcurve function in Matlab, and 95% confidence intervals were estimated using 1000 boot-strap replicas. HFO frequency, power and duration values were fit with generalized linear mixed-effects models (GLMMs) in Matlab with patient as the random-effects term, and SOZ and location as fixed-effects predictors. Violin plots, that are like box plots but also show the smoothed probability density values of the data at different values, were generated in Matlab. All graph theoretical measures were calculated using the Brain Connectivity Toolbox (https://sites.google.com/site/bctnet/)^65^(Supplemental Methods). The adjacency matrix for the distance networks were calculated as the Euclidian distance (mm) between every electrode contact (*i.e.* node) using normalized MNI coordinates. The adjacency matrix for the rate-distance networks was calculated by the average rate (/min) of the events recorded by two respective nodes multiplied by the Euclidian distance (mm) between these nodes. The adjacency matrix for the mutual information (MI) networks were calculated using event “spike trains” defined by the onset times of each event and then calculating MI between nodes using the adaptive partition using inter-spike intervals MI estimator^67^. We compared the MI values between nodes, the local efficiency of nodes, and the strength of nodes using GLMMs with patient as the random-effects term, and SOZ and responder/non-responder, and the interaction between SOZ and responder/non-responder as fixed-effects predictors. Networks were visualized using BrainNet viewer^68^. Principal component analysis, and unpaired t-tests were performed in Matlab (Supplemental Methods). The SVM was trained and tested using Matlab functions fitcsvm and predict. The model was trained after normalizing the data and using a Radial Basis Function kernel that was automatically scaled. We defined as true positive a patient who was correctly classified as a non-responder by the SVM. Estimates of the 95% confidence intervals (CI) used the binomial method.

## Results

### Patient characteristics

A total of 35 patients with medically refractory epilepsy were included in the exploratory portion of the study and 69% of them were male (Table S1). The patients had diverse etiologies of their epilepsy (Table S1) with 13 of the 35 who had non-lesional epilepsy on MRI. Overall, the SOZ was localized to the mesial temporal lobe in 4 patients, lateral temporal lobe in 2, mesial and lateral temporal lobe in 7, temporal lobe and extra-temporal areas in 12, and extra-temporal in the remaining 10 (Table S1).

A total of 21 of the 35 patients had a resection or thermal ablation performed, 8 patients were implanted with a responsive neurostimulation device (Neuropace™, RNS), 1 patient had resection and RNS, 1 patient received a vagal nerve stimulator due to multifocal seizures, 1 patient had a corpus callosotomy due to bilateral multifocal seizure onsets, 2 patients were not offered any surgical intervention due to multifocal seizure onsets, and 1 patient declined RNS (Table S1).

Of the 30 patients who had a resection, ablation, and/or were implanted with an RNS device 19 had a reduction in seizures corresponding with Engel Class I though IVa outcome and were classified as “responders” in this study. Nine patients had no change in seizure frequency (Engel IVb or IVc), including one who died of definite sudden unexpected death in epilepsy (SUDEP) 6 weeks after surgery, and were classified as “non-responders”. Two of the 30 patients were lost to follow up prior to one year, and these patients were not classified as responders or non-responders. We separately examined the 4 patients who were not offered resective or ablation surgery or RNS due to diffuse multifocal seizures (Table S1).

### Differences between surgical responders and non-responders in SOZ classification accuracy using HFO and spike rates

According to the EZ theory, if the EZ and SOZ are discordant, then a resection that targets the SOZ should result in little to no improvement in post-operative seizure outcome (Figure 1A). We used rates of HFO as a proxy for the EZ^24, 26, 27, 30, 31, 69, 70^ to test the hypothesis that there is less overlap of EZ with the SOZ in non-responders than in responders. We calculated receiver operating characteristic (ROC) curves for each HFO subtype and sharp-spikes (Figure 1B) rates in classifying the SOZ for each of the patient cohorts (Figure 2A-E). Results show the area under the ROC curve (AUC) for SOZ classification was not significantly different for the HFO subtypes or sharp-spikes rates between responders and non-responders (bootstrapping, n=1000 surrogates, p>0.05, Figure 2B,C). The responders with a seizure free outcome had AUCs for SOZ classification that trended larger than the non-responders, but only the AUC of FR on spikes (fRonS) approached significance (p=0.05, Figure 2D). In the four patients with multifocal seizures who did not receive resection/ablation or a RNS device (*i.e.* no resection/RNS group), the AUC for fast ripples (FR) on oscillations (fRonO) and fRonS trended smaller than in the responders and non-responders (p=0.05, Figure 2E). Since FR are thought to be a biomarker of the EZ, these results suggest the EZ overlaps with the SOZ as much in non-responders as it does in responders, and both groups have greater overlap of EZ with SOZ than patients with diffuse multifocal seizures.

**Figure 2:**
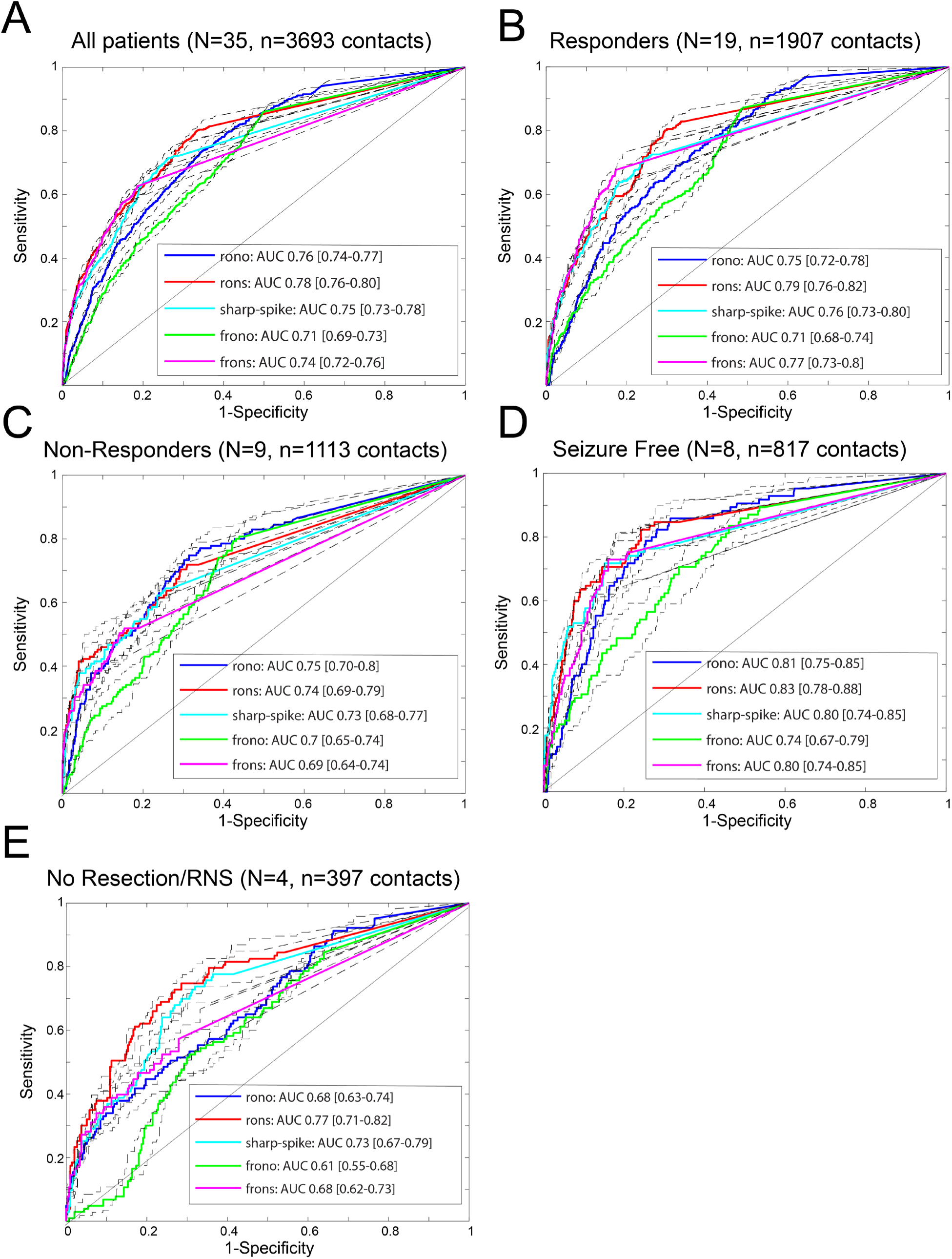
Receiver operator characteristic (ROC) curves for seizure onset zone classification using HFO subtype and sharp-spike rates for the different patient cohorts, (A) all patients, (B) responders, (C) non-responders, (D) seizure free responders, (E) no resection/RNS. Abbreviations (rono: ripples on oscillations, rons: ripples on spikes, frono: fast ripples on oscillations, frons: fast ripples on spikes, AUC: area under the ROC curve). Dashed lines and brackets indicate 95% confidence intervals calculated using bootstrapping (n=1000 surrogates). The AUC for fRonO trended lower in the no resection/RNS group (bootstrapping, p=0.05). The AUC for fRonS trended higher in the seizure free group compared to non-responders (bootstrapping, p=0.05), and was significantly greater than the no resection/RNS group (bootstrapping, p<0.05).

Classification accuracy for the SOZ has been found to vary by HFO subtype and neuroanatomic region^71–73^. In the whole brain at high specificities, rates of HFO on spikes and sharp-spikes were more sensitive for the SOZ than rates of HFO on oscillations among the 35 patients (p<0.05, Figure 2A). In accord with prior studies^72^, we found that sharp-spike rates were comparable to HFOs for classifying the SOZ (p>0.05, Figure 2A-D). In the non-responders and no resection/RNS groups, ripples on oscillation (RonO) rates were equally sensitive compared to HFO on spike rates, but fRonO rates were less sensitive (p<0.05, Figure 2C,D). When we examined SOZ classification accuracy by neuroanatomic region (Supplemental Figure 1), we found that HFO and sharp-spike rates trended better in classifying the SOZ in the frontal lobe neocortex and limbic regions (i.e., cingulate gyrus, perirhinal gyrus, para-hippocampal gyrus). In the hippocampus, fRonO rates performed relatively well for classifying the SOZ, and RonO rates performed at chance^21^ (p<0.05). The lowest SOZ classification accuracy using the sharp spike or HFO rates was in the occipital lobe neocortex^19^. Because the EZ does not always equate with the SOZ^3^, these differences in SOZ classification accuracy do not imply superior accuracy for predicting post-operative seizure outcome^32^.

### Differences in fast ripple spectral frequency between surgical responders and non-responders

Since the HFO rates in the responders and non-responders classified the SOZ with equal accuracy, we reasoned there could be a subset of HFO with unique properties that are more strongly associated with the SOZ. Using a generalized linear mixed-effects model (GLMM) results show in the responders, the peak spectral frequency of fRonO was higher in the SOZ than in the non-SOZ (p<1e-5, Table 1, Figure 3A). By contrast, in the non-responders, the peak frequency of fRonO was lower in the SOZ than the non-SOZ (p<1e-5, Table 1, Figure 3A). In the no resection/RNS group there was no difference in fRonO spectral frequency between the SOZ and non-SOZ (Table 1, Figure 3A). fRonS occurred far less often than fRonO and we did not find significant differences in fRonS peak spectral frequency in the SOZ than non-SOZ for responders or non-responders (Table 1, Figure 3B). In the case of the no resection/RNS group, fRonS peak frequency was significantly higher in the non-SOZ (p<1e-5, Table 1, Figure 3B).

**Figure 3:**
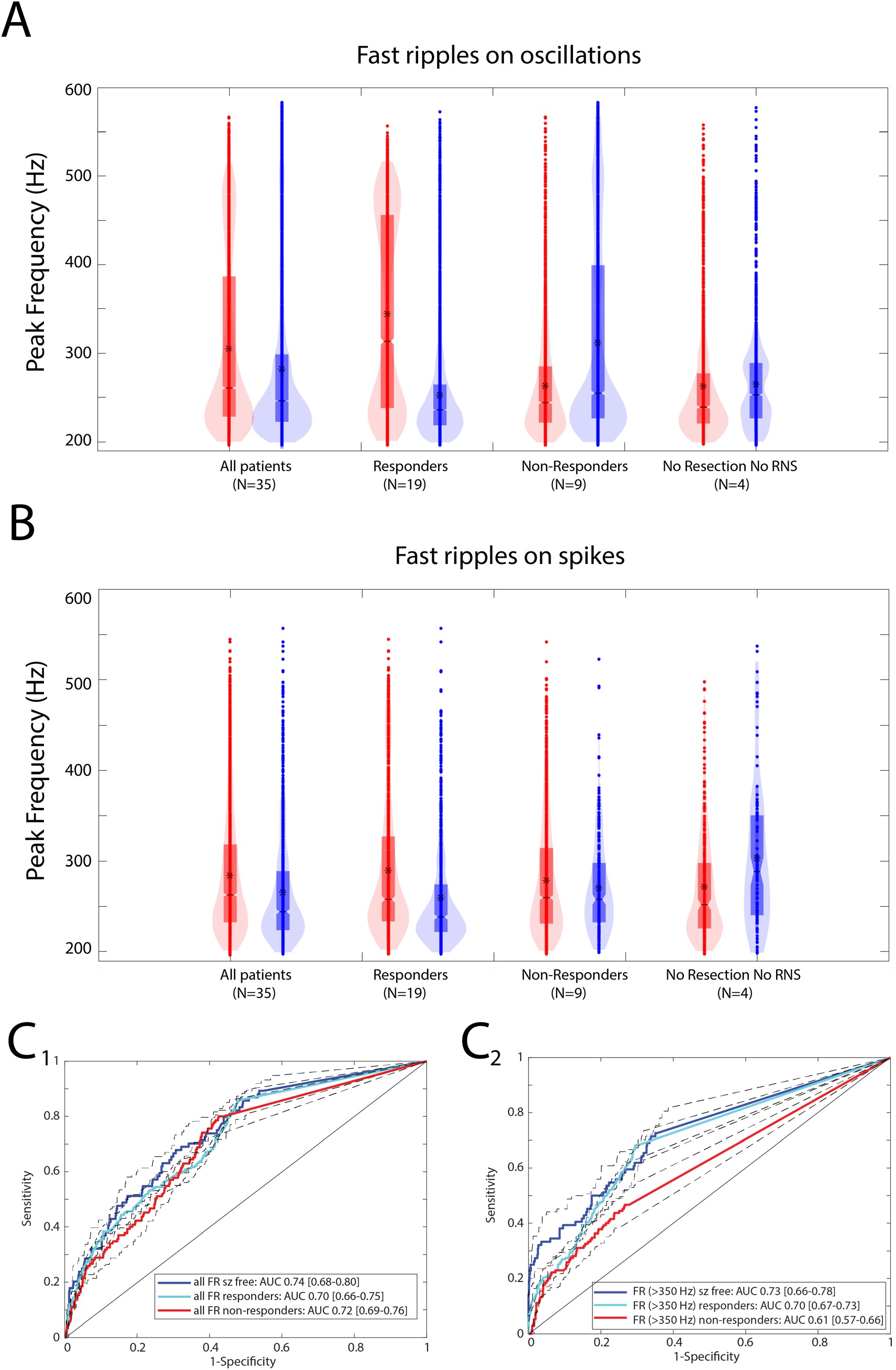
Fast ripples (FR) with a higher spectral content are better markers of epileptogenic brain regions. Violin plots of FRon oscillation (fRonO, A) and FRon spike (fRonS, B) peak spectral frequency in the seizure onset zone (SOZ, red) and non-SOZ (blue) in all patients, responders, non-responders, and patients not offered resection or RNS. Asterisk indicates mean. In the responders, the peak spectral frequency of fRonO was higher in the SOZ than in the non- SOZ (generalized linear mixed-effects model, GLMM, p<1e-5). In the non-responders, the peak spectral frequency of fRonO was lower in the SOZ than the non-SOZ (GLMM, p<1e-5). In the no resection/RNS group, fRonS peak spectral frequency was significantly higher in the non-SOZ (GLMM, p<1e-5). (C) Receiver operator characteristic (ROC) curves for seizure onset zone classification using the rate of all fast ripples, including fRonO and fRonS irrespective of frequency (C1), and higher-frequency fast ripples (fRonO and fRonS >350 Hz) for the different patient cohorts. The area under the ROC curve of FR (>350 Hz) rates was significantly different in responders compared to non-responders (bootstrapping, p<0.05). Dashed lines and brackets indicate 95% confidence intervals calculated using bootstrapping (n=1000 surrogates).

**Table 1:** Results of generalized linear mixed-effects models fitting fRonO frequency in the different patient cohorts. The random-effect term was the patient, the fixed effects were the SOZ, and the location of the electrode. Brackets indicate 95% confidence interval. Abbreviations n.s. not significant.

| Response Variable | Intercept estimate | Intercept p-value | SOZ estimate | SOZ p-value | Location estimate | Location p-value | d.f. |
| --- | --- | --- | --- | --- | --- | --- | --- |
| All patients fRonO freq | 5.556<br>[5.51 5.60] | <1e-5 | -0.081<br>[-.086 -.076] | <1e-5 | 0.016<br>[.014 .017] | <1e-5 | 41168 |
| Responders fRonO freq | 5.470<br>[5.41 5.52] | <1e-5 | 0.178<br>[.170 .187] | <1e-5 | 0.012<br>[.010 .013] | <1e-5 | 17619 |
| Non-Responders fRonO freq | 5.646<br>[5.53 5.76] | <1e-5 | -0.352<br>[-.362 -.343] | <1e-5 | 0.010<br>[.008 .011] | <1e-5 | 15283 |
| No resction or RNS fRonO freq | 5.523<br>[5.47 5.58] | <1e-5 | -0.004<br>[-.015 .008] | n.s. | 0.014<br>[.012 .017] | <1e-5 | 6022 |
| All patients fRonS freq | 5.591<br>[5.54 5.64] | <1e-5 | -0.021<br>[-.039 -.003] | 0.02 | 0.009<br>[.005 .012] | <1e-5 | 6302 |
| Responders fRonS freq | 5.522<br>[5.45 5.60] | <1e-5 | 0.005<br>[-.024 .030] | n.s. | 0.0175<br>[.013 .023] | <1e-5 | 2430 |
| Non-Responders fRonS freq | 5.613<br>[5.56 5.67] | <1e-5 | -0.008<br>[-.036 .020] | n.s. | 0.005<br>[-.0005 .01] | n.s. | 2649 |
| No resction or RNS fRonS freq | 5.768<br>[5.69 5.85] | <1e-5 | -0.097<br>[-.14 -0.05] | <1e-5 | -0.018<br>[-.031 -.005] | <1e-3 | 465 |

We examined the effects of neuroanatomic location on fRonO and fRonS frequency in the SOZ and non-SOZ. We found that in almost all patient groups, neuroanatomic location significantly influenced the peak spectral frequency of fRonO and fRonS (p<1e-5, Table 1).

Mean fRonO peak frequency was higher in the SOZ than the non-SOZ for frontal and temporal lobe neocortex and limbic regions (Supplemental Figure 2A). Mean fRonS peak frequency was higher in the SOZ than the non-SOZ only in frontal lobe neocortex (Supplemental Figure 2B).To determine whether FR with a higher spectral frequency were better biomarkers of epileptogenic regions we compared, in the different patient cohorts, the AUCs for SOZ classification of FR rates. In calculating these rates, we combined fRonO and fRonS events (Figure 3C1) and examined FR events with a peak spectral frequency > 350 Hz (Figure 3C2). We found that the AUC for SOZ classification with FR >350 Hz was significantly higher in the responders as compared to non-responders (bootstrapping, n=1000 surrogates, p<0.05).

Next, we examined whether there were differences in ripple peak spectral frequency, HFO power, or duration between responders and non-responders. GLMMs for these parameters indicated that ripple on spike (RonS) peak spectral frequency was slightly lower in the SOZ for responders, but slightly higher in the SOZ for non-responders (p<1e-5, Supplemental Table 2). RonO peak power was slightly increased in the SOZ for responders, but the effect size was significantly larger for non-responders (p<1e-5, Supplemental Table 3). In contrast, for RonS peak power the effect size was significantly larger for the responders (p<1e-5, Supplemental Table 3). fRonO peak power was slightly decreased in the SOZ of responders (p=0.006) but was increased in the SOZ of non-responders (p<1e-5, Supplemental Table 3). fRonS peak power was significantly increased in the SOZ of responders (p<1e-5), but not non-responders (Supplemental Table 3). Regarding HFO duration, we found no clear effects of the SOZ that were related to response to surgery (Supplemental Table 4).

### Differences in the fast ripple (> 350 Hz) rate-distance networks of surgical non-responders

Since we found no difference in classification accuracy of the SOZ using HFO rates between responders and non-responders, except when using FR > 350 Hz, we considered whether the spatial distribution of HFO-generating sites, specifically fRonO and fRonS with peak spectral frequency >350 Hz, would discriminate responders from non-responders. For this analysis we computed the radial distance of the network formed from electrodes that recorded at least a single FR > 350 Hz. This measurement was made in Euclidian space and was independent of neuroanatomical boundaries and white matter tracts. We selected this criterion because prior research has shown that the occurrence of one FR can predict seizure recurrence^24, 26, 69^. Since the no resection/RNS patients were considered non-responders *a priori* they were included in the non-responder’s group to permit binary classification. We excluded non-responder IO010 because we were unable to characterize the complete network due to iEEG contamination on 40% of the electrode contacts. Analysis showed for most patients the radius of the FR network (Figure 4B) was larger than the radius of the SOZ (Figure 4A,5A). One outlier was patient 4100 who had no fast ripples > 350 Hz. This patient reported an Engel IVb outcome and underwent prolonged post-operative scalp EEG monitoring. After withdrawing anti-seizure drugs, no sharp-waves, spikes, or seizures were recorded. Among all the patients, the radius of SOZ and radius of FR distance networks performed sub-optimally at classifying non-responders from responders (Figure 5A2). In comparison to the responders, the radius of the SOZ and FR distance networks was larger only for the non-responders who were not selected for resection/RNS due to multi- focal onsets, or patients with bilateral SOZ sites who underwent resection and had an Engel IVb/c outcome (Figure 5A1).

**Figure 4:**
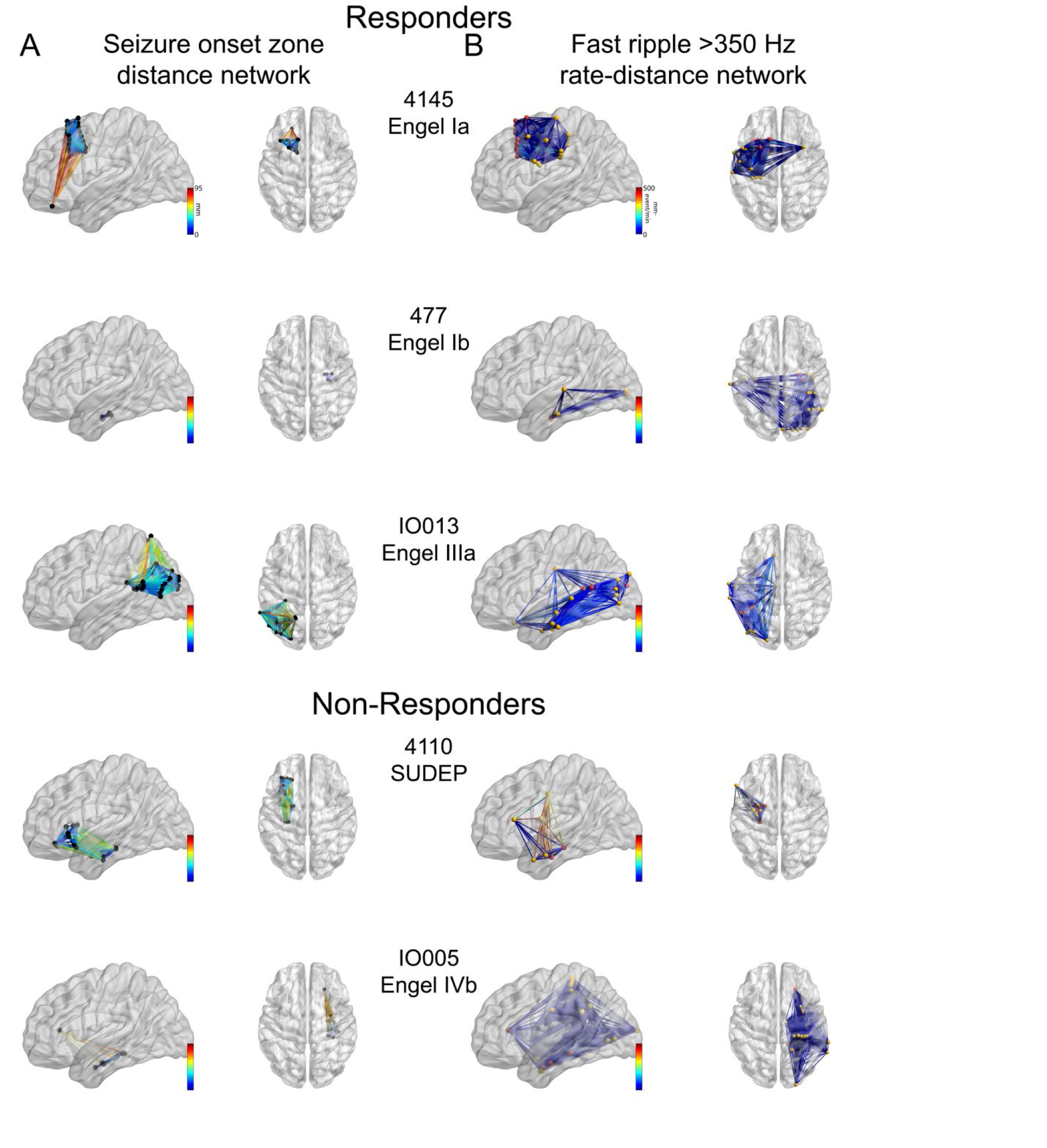
Glass brain renderings of the (A) SOZ distance networks and the (B) FR (> 350 Hz) rate-distance networks for three representative responders (top) and two non-responders (bottom). (A) The edge color corresponds to the geometric distance (mm) between electrode contacts inside the SOZ. (B) The electrode contacts in the SOZ are colored red and those in the non-SOZ yellow. The edge color corresponds to the geometric distance multiplied by the average FR rate between the two electrodes. The FR distance networks (not shown) can be inferred from (B) since the node locations are identical, but the edge weights are calculated as the Euclidian distance between the nodes alone (not shown).

**Figure 5:**
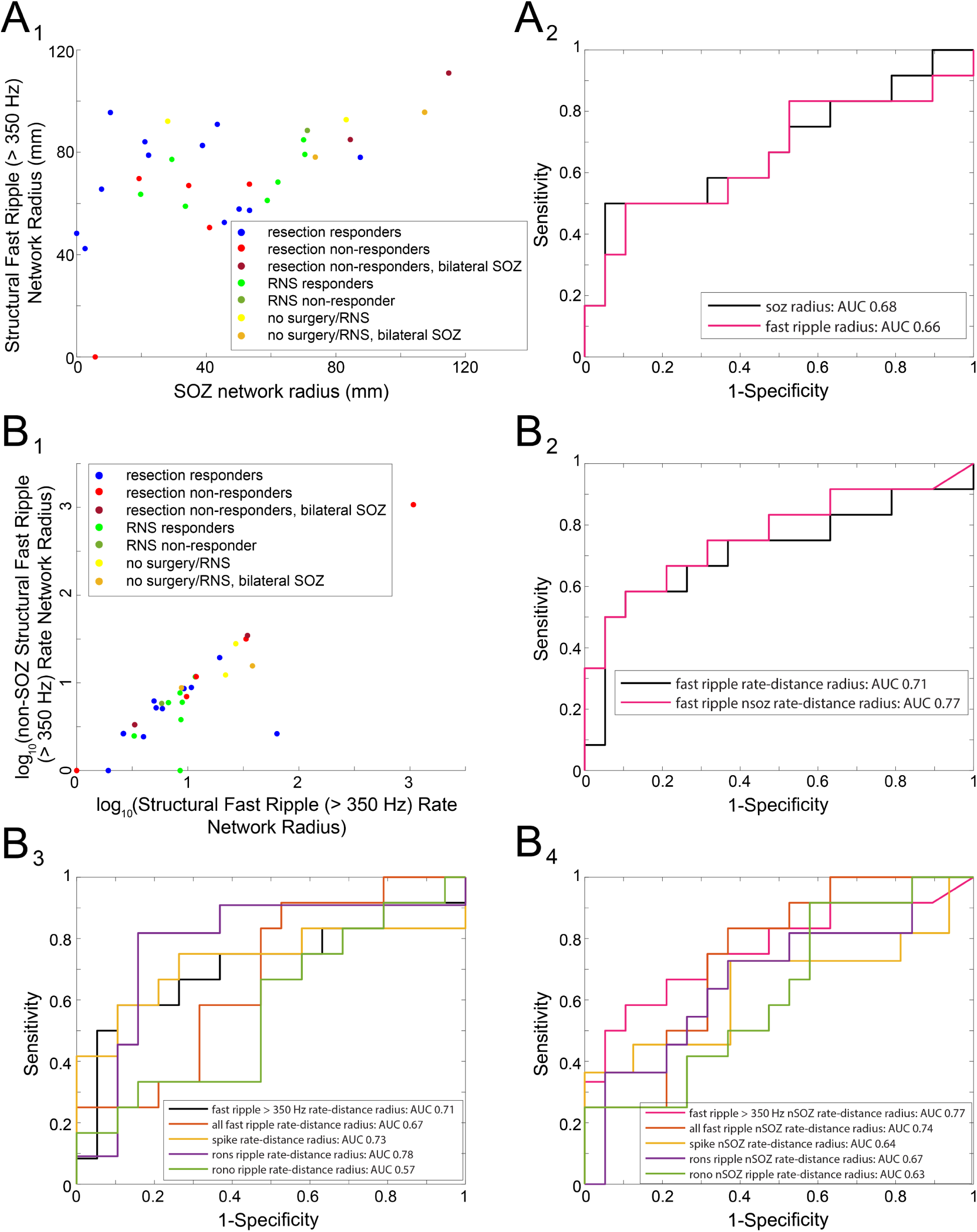
FR (>350 Hz) rate-distance network for each patient group and classification accuracy of non-responders and no resection/RNS patients. (A1) Scatterplot of the radius of the SOZ and the radius of the FR network for the 31 patients (A2) Receiver operating characteristic (ROC) curves of the radius of the SOZ network (black) and radius of the FR network (magenta) for classifying non-responders. (B1) Scatterplot of the log transformed radius of the FR rate-distance network, and the log transformed radius of the corresponding non-SOZ networks. (B2) ROC curves of the radius of the FR rate-distance networks (black) and corresponding non-SOZ networks (magenta). After bootstrapping (n=1000 surrogates, not shown), the AUC for the FR non-SOZ rate distance radius was 0.77 [95% confidence interval (CI) 0.56-0.98]. (B3) ROC curves of the radius of the rate-distance networks of the different biomarkers. (B4) ROC curves of the radius of the corresponding non-SOZ networks of the different biomarkers.

Since many responders had FR sites outside the SOZ, which were probably not resected, and there were small differences in the spatial distribution of FR generating sites between responders and non-responders, weighting the edges of the distance graphs by the average FR (> 350 Hz) rates from the two electrodes could improve the sensitivity in discrimination patient’s response to surgery. We constructed two rate-distance graphs for the patients. The first graph included all electrodes with at least one FR (> 350 Hz), while the second graph was a subset of the first and included only electrodes in the non-SOZ. We found that these rate-distance networks, and the radius of these networks, performed better at classifying responders with a smaller radius from non-responders (Figure 4B,5B1,B2). The radius of the rate-distance network constructed from electrodes in the non-SOZ had an AUC of 0.77[95% confidence interval (CI) 0.56-0.98] for classifying non-responders (Figure 5B2). To verify fast ripples > 350 Hz were superior to other HFO subtypes in differentiating responders from non-responders, we constructed the same rate-distance networks using all FR as well as other HFOs. This analysis showed the radius of RonS rate-distance networks was best in classifying responder from non-responders but had low sensitivity at high specificities (Figure 5B3). In the case of the rate-distance networks for the non-SOZ, the radius of the rate-distance using FR >350 Hz performed the best in classifying responders and non-responders (Figure 5B4).

### Differences in the fast ripple (> 350 Hz) mutual information networks of surgical non-responders, and classification of non-responders using machine learning

Long-range synchronization^74^ and propagation^34^ of HFOs is known to occur, and we used mutual information (MI) to assess the timing between FR (>350 Hz) recorded from every pair of electrode contacts. Constructing FR networks using MI provides a measure of how much information in the occurrence of FR at one electrode tells us about FR on another electrode. In this analysis, we were able to construct FR MI networks in 13 of the 19 responders, and 9 of the 12 non-responders. In the others, FR events occurred too infrequently. FR networks with nodes in the non-SOZ were found in only 11 of the responders and 8 of the non-responders.

Across patients, the topology of FR MI networks were heterogenous and within the responder and non-responder cohorts the topology remained inconsistent (Figure 6). To identify network features that distinguished responders from non-responders we applied GLMMs to MI, local efficiency, and nodal strength that accounted for within-patient effects. We first examined the MI between SOZ:SOZ, SOZ:non-SOZ, and non-SOZ:non-SOZ nodes (Figure 7B). We found a trend that the MI value of the edge depended on whether the respective nodes were in the SOZ (p=.09), but no difference between responders and non-responders (Table S5). Our first global measure, the characteristic path length, which is the average shortest path length in the correlational FR MI network, was longer in some non-responders (e.g., see 4110 in Figure 6), but some responders also exhibited a long path length too (e.g., see 4145 in Figure 6t-test, p=0.47, n=8 and 11, Figure 7A, Figure S3).

**Figure 6:**
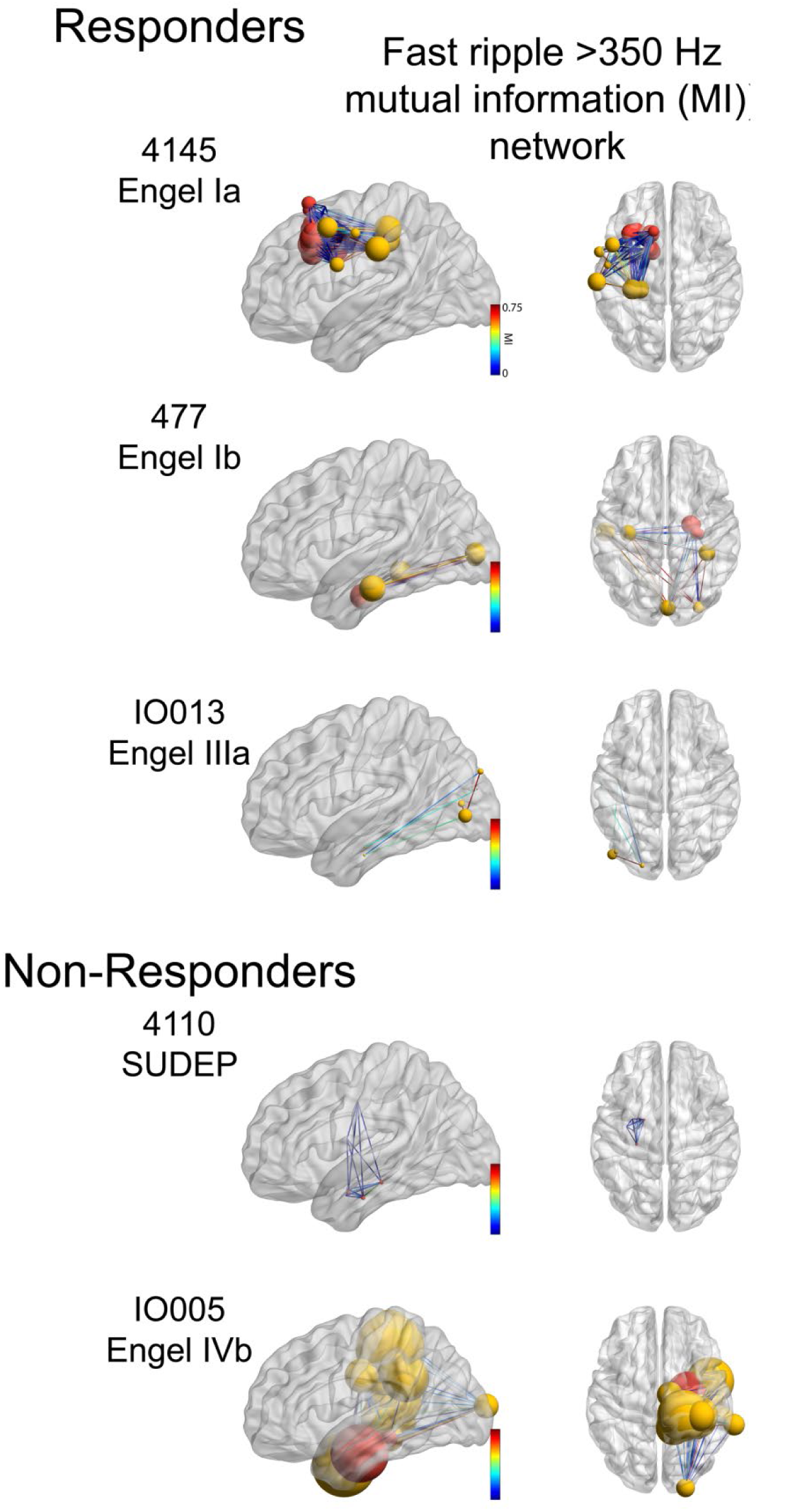
Glass brain renderings of the fast ripple (> 350 Hz) mutual information networks defined by mutual information between fast ripple “spike trains” recorded from paired electrode contacts. Nodes in the SOZ are colored red, nodes in the non-SOZ are colored yellow. The size of the node corresponds to the node strength. The edge color corresponds to the mutual information value between nodes.

**Figure 7:**
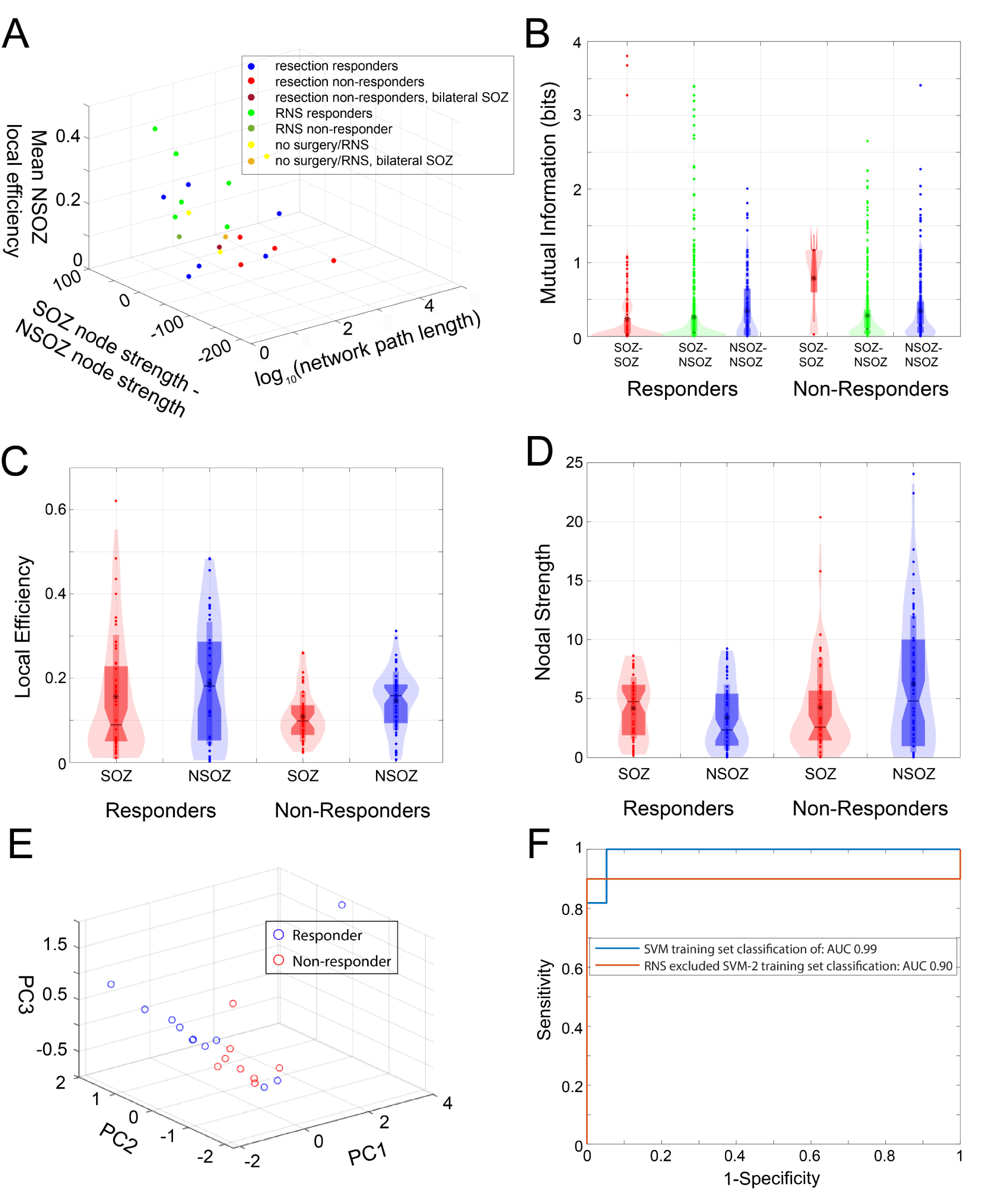
Graph theoretical measures of the fast ripple (FR > 350 Hz) mutual information (MI) networks improves the discrimination of non-responders. (A) Three-dimensional scatter plot of the FR (> 350 Hz) MI global graph theoretical measures (three univariate t-tests, p>0.05). (B) Violin plot of MI computed for SOZ-SOZ edges (red), SOZ-non SOZ (NSOZ) edges (green), NSOZ-NSOZ edges (blue) across all the responder and non-responder patients (Generalized linear mixed-effects model, GLMM, p>0.05). (C) Local efficiency of SOZ (red) and NSOZ (blue) nodes across responder and non-responder patients (GLMM, p>0.05). (D) Nodal strength of SOZ (red) and NSOZ (blue) nodes across responder and non-responder patients. The location of the node within the SOZ significantly influenced nodal strength (GLMM, p<0.05), (E) Three- dimensional scatter plot of the principal component (PC) scores derived from principal component analysis of the three global measures (A) from all the patients. The PC2 score was significantly different in the responders and non-responders (t-test, p=0.03), the PC1 and PC3 scores were not significantly different (t-test, p>0.05). (F, blue) The receiver operating characteristic curve (ROC) for non-responder classification in the exploratory dataset (n= 19 responders, n=11 non-responders/no resection or RNS) by the support vector machine (SVM) trained using the SOZ, and FR (> 350 Hz) distance, rate-distance, and three MI global metrics derived from these patients. (B, red) The ROC curve for non-responder classification in the exploratory dataset without RNS implant only patients (n = 12 responders, n = 10 non- responders/no resection or RNS) using the SVM trained on the SOZ, and fast ripple (> 350 Hz) metrics from these patients. Abbreviations: AUC: area under the ROC curve.

We next calculated the local efficiency, which is the inverse shortest path length in the network computed on the neighborhood of the node, for all the connected nodes for each patient (Figure 7C). We found a trend that the local efficiency is affected by whether the node was in the SOZ (p=.12, Table S5), and the patient’s status as a responder/non-responder (p=.10, Table S5). Our second global measure, the mean local efficiency of the non-SOZ nodes, was increased in many responders compared to non-responders (t-test, p=0.07, Figure 7A, Figure S3).

We next examined the nodal strength for all the connected nodes in all the patients (Figure 7D). We found that the location of the node within the SOZ significantly influenced nodal strength (p<0.05, Table S5), and that the interaction of the location of the node within the SOZ and the patient’s status as a responder/non-responder trended towards significance (p=.12, Table S5). We computed our third global measure, the difference in nodal strength among summed non-SOZ nodes relative to SOZ nodes and found that it appeared to vary in some of the responders relative to non-responders (t-test, p=0.26, Figure 7A, Figure S3). None of these three global measures calculated using the other HFO types, including all FR irrespective of frequency, could visibly cluster surgical responders away from non-responders (Figure S4).

To account for the variability in the topology of the FR networks we sought to combine the three global measures. A SVM is trained by constructing a hyperplane of the multidimensional data for classification purposes^75^, to test if the three measures would contribute to an accurate hyperplane, we applied principal component analysis (PCA) to the three global measures (Figure 7E). The scores of principal component 1 (PC1) were not significantly different between responders and non-responders (t-test, p=0.35), but PC2 scores were significantly different (t-test, p=0.03), and PC3 scores trended towards significance (t-test, p=0.10). Thus, the variance in the three measures combined explained by PC2 indicate that the three measures would serve as useful factors for the SVM to construct an accurate hyperplane.

We next trained the SVM using the three global measures of the FR (> 350 Hz) MI networks, as well as the distance, rate-distance radii calculated in the exploratory dataset patients. The trained SVM was tested to classify non-responders from responders in a distinct test of 13 patients who had resection, unilateral RNS, or were not deemed resection/RNS candidates due to multifocal seizures (Table S6). We first tested the accuracy of the trained SVM on the patients in the exploratory dataset (Figure 7F, Table S7). We included SOZ node radius in the SVM, because we also sought to identify patients not offered surgery based on iEEG monitoring. The false negative patient 4100 was excluded from the training set. We found that the SVM (SVM-1) could classify the non-responders in the training set with 99% accuracy (Figure 7B, Table S7). However, in the test dataset, the sensitivity of SVM-1 using a threshold SVM score of 0.5 was 0.25 [95% confidence interval (CI) 0.05–0.53], the specificity was 1.0 [95% CI 0.75-1.0], the positive predictive value (PPV) was 1.0 [95% CI 0.75-1], the negative predictive value (NPV) was 0.75 [95% CI .39-.91], and the accuracy was 0.77 [95% CI .55-.98] (Table S6 & S8). Despite the low sensitivity, the SVM-1 score was significantly different between the responders and non-responders in the test set (t-test, n=9,4, p=0.002). Since the sensitivity of SVM-1 was low, and all but one of the RNS patients was classified as a responder, we hypothesized that excluding the patients treated with RNS alone from the training set would improve the sensitivity of classifying of non-responders in the test set (Figure 7F). This SVM (SVM-2), which may have limited generalizability because it excluded patients in the training set, classified non-responders in the test set with a sensitivity of 0.75 [95% CI 0.39-0.9], a specificity of 1.0 [95% CI 0.75-1], a PPV of 1.0 [95% CI 0.75-1], a NPV of 0.9 [95% CI 0.75-1], and accuracy of 0.92 [95% CI 0.75-1] at a score threshold of 0.5 (Fig 7F, Table S6 & S8).

## Discussion

Substantial evidence has accumulated supporting the epileptic network hypothesis in epileptogenesis and ictogenesis^4–10^, but the neurophysiological mechanisms that underlie these networks are not well understood, particularly in the context of prior established frameworks^76, 77^. Using bilateral invasive EEG recordings in a large and diverse cohort, we provide evidence that shows in patients without post-operative reduction in seizure frequency or in patients deemed poor candidates for resection or RNS, FR can form a decentralized network consisting of widespread, hyperexcitable FR-generating nodes. FR networks are believed to be a mechanism for ictogenesis and, although we did not characterize the resection volume, our results imply that such FR networks may continue to generate seizures even after a portion has been resected or disconnected. These results also suggest that in patients who are seizure-free, epileptogenic regions that meet the criteria of an EZ might also be a focal FR network where critical hubs have been removed but some remote sites remain (Figure 1A).In support of this notion intraoperative recordings of staged resections have shown that FR sites present in the initial recording, disappear in subsequent recording following a resection of the presumed hubs of the FR network^26^.

In contrast to the numerous studies investigating FR as a biomarker of the EZ in post- operative seizure free patients, we postulated that a corollary of the EZ hypothesis is a larger discordance between FR-generating sites and the SOZ(s) in non-responders (Figure 1A), which does not necessitate a single EZ region. We found ripple and FR rates are equally accurate for defining the SOZ(s) in seizure free responders, all responders, and non-responders, suggesting that at least a portion of the putative EZ was resected or disconnected in the non-responders. The EZ theory states that the EZ is indispensable for seizure generation^1, 2^, so it is unexpected that patients would not experience some reduction in seizure frequency after surgery. However, in support of the EZ hypothesis, the rate of FR (>350 Hz), and fRonS in seizure free patients, better classified the SOZ in responders than non-responders suggesting that the SOZ may have been more discordant with the EZ in the non-responders at the group level.

At the individual level, an SVM trained on the graph theoretical metrics of the FR networks was able to accurately classify non-responders from responders, indicating an epileptic network-guided approach could be useful in planning epilepsy surgery.

Our findings add to a growing literature that has used the epileptic network hypothesis and graph theory ^4–10^ to explore epileptogenesis^18, 20^ and ictogenesis.^78–80^ ^18, 2278–804–10^ Prior studies examining static and dynamic correlational iEEG networks in epilepsy surgery patients have been critical of the utility of using HFOs for improving epilepsy outcomes^37, 38, 81^, but our study demonstrates the feasibility and utility of constructing FR networks that could also improve seizure outcome. Thus, it is important to understand the mechanisms responsible for FR networks and how to localize these networks to help plan epilepsy surgery^82, 83^.

### Neurophysiological Mechanisms Underlying Fast Ripple Networks

The FR rate-distance and MI networks distinguished non-responders more accurately when FR <350 Hz were excluded. This is because in responders, FR > 350 Hz were found more often in the SOZ and in non-responders, they were found more often in the non-SOZ. Very fast and ultrafast ripples with spectral frequency > 500 Hz have been previously reported as specific biomarkers of the EZ^84–86^. Modeling studies have shown that FR spectral frequency is partially determined by the reversal potential of GABAergic synapses, with a more depolarizing reversal potential producing higher frequency FR^87^. Our results suggest including spectral frequency of FR in the analysis, and ultimately understanding the mechanisms that generate FR, are important. Higher-frequency FR appears to more strongly associate with epileptogenic regions.

In the patients in which FR MI networks could not be characterized, the classification of non-responders relied entirely on the distance and rate-distance networks. The rate-distance network is an intuitive measure of the spatial geometry of FR-generating sites weighted by FR rate and the radius gauges how widespread and active the fast ripple generating contact sites are relative to each other. This network does not consider anatomical or functional connectivity between sites. In contrast, the MI networks used mutual information to compare FR timing between sites. This measurement is sensitive to FR synchrony and propagation, and is thus a measure of FR functional connectivity. In humans, FR can propagate over short distances (average of 16 mm) in channels inside the SOZ,^34^ though longer distances in FR synchrony are possible^74^.

Evidence for FR networks during epileptogenesis is found in the unilateral hippocampal kainic acid model of mesial temporal lobe epilepsy^88, 89^. The injected hippocampus generates FR and other epileptic activity and during delta EEG rhythms, drives and synchronizes FR in frontal neocortex^89^. In our study, FR were detected during non-REM sleep, and it is possible that coupling with delta rhythms could similarly drive remote FR and thus contributed to the MI of FR.

We found that the characteristic path length was sometimes longer in the FR MI networks of non-responders. Although, the MI values of the individual edges were similar. While an increase in the number of nodes generating FR alone could have contributed to the longer path length, we interpret the difference as an indication of asynchrony between the FR generating sites in the non-responders. The MI values between nodes were too low for propagation of FR between nodes to account for the longer path length. Thus, since the spatial sampling of stereo EEG is intrinsically limited, an increase in the number of nodes implies an increase in asynchrony.

We also found that many of the non-responders had a lower mean local efficiency in the non-SOZ nodes, but a greater number of high strength non-SOZ nodes relative to SOZ nodes. These results suggest in the non-SOZ of non-responders, FR-generating sites were more numerous, more interconnected, and FR were more asynchronous than in the responders.

FR are thought to be generated by action potentials from clusters of pathologically interconnected neurons (PIN)^18^. The PIN cluster hypothesis proposes PIN clusters can act as internal kindling generators that potentiates synaptic connections in target areas and recruits additional structures^17^. Our FR rate-distance and MI networks results are consistent with this hypothesis and indicate in non-responders, PIN clusters are widespread. In patients with advanced disease, one or more PIN clusters may form hubs connected with modules, which results in greater autonomy and asynchrony in the FR network, especially outside the SOZ. Alternatively, if FR networks are a mechanism for inhibitory surround,^90–92^ then one would expect the strength and extent of the FR network correlates with fewer seizures after surgery, rather than current results of no change in seizure frequency. Moreover, while individual seizures may begin in an ictal core region smaller than the SOZ^90–94^, resecting the core alone may not consistently reduce seizures if different nodes of a FR network can generate seizures.

### Fast ripple network measures and selection and prediction of surgical treatment and outcome

Few studies have found criteria that identify potential surgical non-responders based on the pre-surgical iEEG evaluation^14, 95–97^. RNS is not recommended if more than two independent SOZs are identified^41^. Surgical non-responders experience substantial morbidity and mortality and represent at least 15-20% of epilepsy surgery patients^95, 96, 98^. If these patients were identified accurately, they could be potentially treated with deep brain stimulation or vagal nerve stimulation instead of resection, which would reduce morbidity and improve seizure outcome after surgery. We found that the FR rate-distance networks discriminated non-responders and patients not offered resection or RNS with moderate accuracy. The rate-distance network that included only nodes in the non-SOZ performed slightly better, perhaps because the SOZ nodes were often at least partially resected or targeted with RNS. Prior studies have demonstrated that surgical non-responders, who also failed a repeat epilepsy surgery, exhibit widespread multi- focal inter-ictal discharges. Also, in these patients, the SOZ identified in the repeat iEEG evaluation was discontiguous with the SOZ identified in the initial iEEG evaluation^14, 95–97^. Like the reported widespread inter-ictal discharges in non-responders of these previous studies, we found widespread high-rate FR in non-responders here as well. Also, our interpretation of a decentralized FR network is consistent with potential multiple, discontinuous SOZs.

When the SVM was trained using all the responders and non-responders in the exploratory dataset it accurately classified the patients in the training set, but its sensitivity for predicting non-responders in the test set was low. The sensitivity improved after excluding the RNS patients from the training set. One potential reason is that the radii of the distance and rate-distance networks were relatively large for the RNS patients, who mostly had bilateral SOZs, and all but one was classified as a responder. Thus, the trained SVM emphasized radius less in the overall score. In contrast, when the RNS patients were excluded, the radius was emphasized more in the score, which was important for identifying the surgical non-responders in the test set. Based on our data, it is unknown whether our graph theoretical measures of FR networks can distinguish RNS non-responders. Future work may seek to characterize the location of the RNS stimulation contacts relative to the FR network nodes to better understand and predict the efficacy of neuromodulation.

While we did not explicitly compare the graph theoretical measures of RNS patients to those patients not offered resection/RNS, we found that the radii of the distance and rate-distance FR networks were typically larger in the patients not offered resection/RNS. This result indicates that the clinical selection criteria of RNS candidates in our study were reflected by differences in the patient’s FR networks.

Our study did not quantify the resected volume because not every patient had a resection, and we assumed that in the patients who had resections, at least a portion of the identified SOZ was resected. A follow up study could measure the resected volume and determine how our SVM approach, in combination with computer-simulated resection^38, 39, 99^, compares with the actual surgery. The SOZ nodes used in the calculation of the current graph theoretical metrics could be substituted with resected nodes in the modified SVM. The SVM would then be retrained and retested, but if the accuracy is sufficient, it could be used prospectively to identify and possibly reduce the number of surgical non-responders.

### Study limitations

An important consideration in this study is seizure outcome determined at time of last follow up. Up to 40-50% of patients who were initially surgical non-responders can show some improvement at five years post-initial operation, but in many cases, this is after a second epilepsy surgery^95^. Also, we also included patients not offered surgery/RNS together in the non-responder group. Although our analysis demonstrated similar neurophysiological features between these groups, the latter could have potentially exhibited a surgical response. Like all iEEG studies during presurgical evaluation, information about the SOZ and HFOs is limited to the number and placement of electrodes, which might not fully characterize the area that generates seizures and HFOs.

## Conclusion

The EZ hypothesis has served as the theoretical foundation for resective and ablative epilepsy surgery for decades. FRs have shown promise as a biomarker of the EZ that could be used as a guide to improve the likelihood of a seizure free outcome. Here we show that, particularly in patients that do not benefit from surgery, FR form correlational networks that can be decentralized, relatively desynchronized, and highly active. Moreover, based on the spatial geometry of FR activity, and graph theoretical properties of the FR correlational networks we could accurately distinguish the patients that would either not respond to surgery or not be offered resection/ablation or RNS. Thus, we conclude, that while the EZ hypothesis has utility towards investigating seizure free outcomes, the epileptic network hypothesis better explains patients who do not respond to surgery. It should be emphasized that these two hypotheses are not mutually exclusive as a discrete EZ in a responder may also be conceptualized as a relatively small FR networks with hubs in SOZ contacts, and in non-responders the “EZ” may be discontiguous with the sites of seizure onset. In the future, characterizing both the rate of FR, and graph theoretical measures of FR networks, could improve the clinical outcomes of medically refractory epilepsy patients undergoing presurgical evaluation.

## Data Availability

All data produced in the present study are available upon reasonable request to the authors.

## Data availability statement

The datasets generated during and/or analyzed during the current study are available from the corresponding author on reasonable request.

## Acknowledgements

The authors would like to thank Mr. Dale and Ted Wyeth, and Mr. Kirk Shattuck for their technical contributions. We would also like to thank Ms. Karishma Kurowski for clerical assistance.

