## supplemental methods and figures for "Graph Theoretical Measures of Fast Ripples Support the Epileptic Network Hypothesis"

**HFO Detection**

HFOs and sharp-spikes were detected in the non-REM sleep iEEG using previously published methods^29,62,63^ implemented in Matlab (Mathworks, Natick, MA, USA). In brief, the HFO detector reduced muscle and electrode artifacts in the iEEG recordings using an independent component analysis (ICA)-based algorithm^63^. After applying this ICA-based method, ripples and fast ripples were detected in the referential and bipolar montage iEEG recordings per contact by utilizing a Hilbert detector, in which a 1,000th order symmetric finite impulse response (FIR) band-pass filter in the (80–600 Hz) band for ripples and (250-600 Hz) band for fast ripples was applied, and (ii) a Hilbert transform was applied to calculate the instantaneous amplitude of this time series according to the analytic signal z(t)

z(t)=a(t)e∧iϕ(t)	(1)

where a(t) is the instantaneous amplitude and ø(t) is the instantaneous phase of z(t). Following the Hilbert transform, the instantaneous HFO amplitude function [a(t)] was smoothed using moving window averaging, the smoothed instantaneous HFO amplitude function was normalized using the mean and standard deviation of the time series, and a statistical threshold defined by the skewness of the normalized time series was used to detect the onset and offset of discrete/potential events.

HFO-like events can arise due to Gibb's phenomenon, i.e., high-pass filtering sharp transients, including epileptiform spikes^64^. To distinguish authentic HFOs from authentic HFOs on EEG spikes or spurious HFO due to filter ringing, we used a algorithm that performed topographic analysis of time-frequency plots for each HFO and defined open- and closed-loop contour groups (Figure 1B)^62^. The algorithm also measured the power, spectral content, duration, onset time, and offset time of each HFO and categorized the HFO as an HFO on oscillation, HFO on spike, or sharp-spike (*i.e.* false HFO)^64^. HFO on oscillation refers to all HFOs that do not coincide with a spike.

**Graph Theoretical Measures**

All graph theoretical measures were calculated using the Brain Connectivity Toolbox (<https://sites.google.com/site/bctnet/>)^65^. For the distance networks, the Euclidian distance was calculated between every electrode contact (*i.e.* node) using the normalized MNI coordinates. For the SOZ distance networks, the distance between non-SOZ:SOZ and non-SOZ:non-SOZ nodes were assigned infinite values. For the fast ripple distance networks, the distance between any two nodes that did not both generate a single fast ripple was assigned infinite values. The radius of these networks was calculated by deriving the minimum eccentricity across all the connected nodes using the charpath function. For the biomarker rate-distance networks the Euclidian distance between nodes was multiplied by the average rate (events/min) of the events recorded by the two respective nodes. For the non-SOZ rate-distance networks, the rate weighted distances between SOZ:SOZ and SOZ:non-SOZ nodes were assigned infinite values. To construct the biomarker mutual information (MI) networks, event “spike trains” were defined using the onset times of each event. Edges were assigned weights using the mutual information between the event spike trains of paired nodes. MI was calculated with the adaptive partition using inter-spike intervals MI estimator (AIMIE)^66^, resulting in weighted directed networks. If the MI was zero or not numeric, then the edge was assigned a weight of zero and an infinite distance. To calculate the characteristic path length of the MI network, we inverted the mutual information values to distance and used the charpath function. To calculate the mean local efficiency of the non-SOZ nodes we applied the updated efficiency_wei function to the MI adjacency matrix. By convention, if a MI network could not be defined, or the MI network had no non-SOZ nodes, the mean non-SOZ local efficiency was 1. To calculate the difference in summed nodal strength of SOZ and non-SOZ nodes we applied the strength_dir function to the MI adjacency matrix.

**Statistical Comparison of MI Network Measures and Principal Component Analysis**

For all the patients with MI networks that included nodes in the NSOZ, the characteristic path length of the MI network, the mean local efficiency of the non-SOZ nodes, and the difference in summed nodal strength of SOZ and non-SOZ nodes was compared between responders and non-responders using the ttest2 function in Matlab. Principal component analysis was applied to these three measures from all patients, irrespective of responder status, using the pca function in Matlab resulting in three principal components. The ttest2 function was used to statistically compare the scores of the principal components between the responders and non-responders.

**Figures**

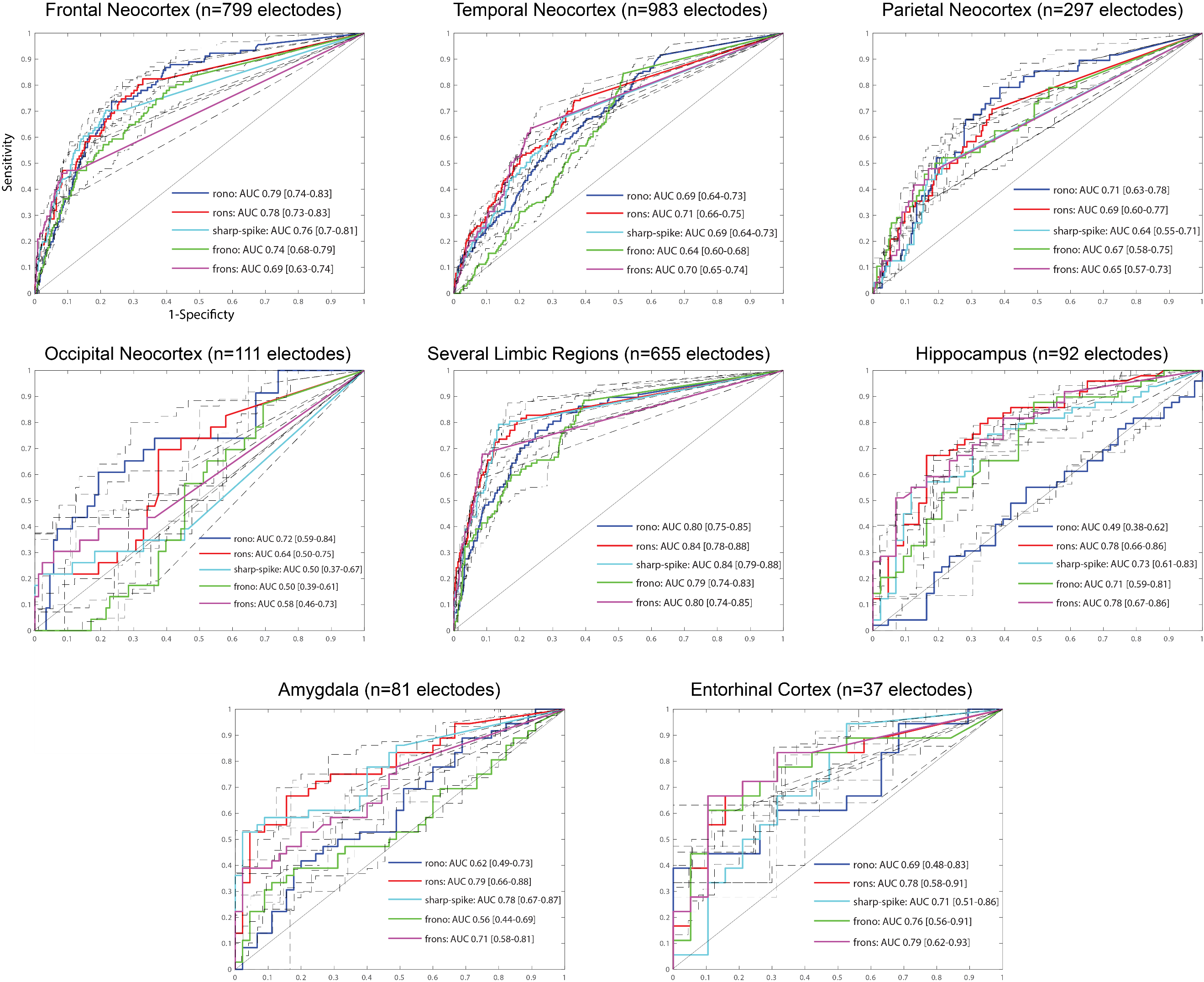

Figure S1: Receiver operator characteristic (ROC) curves for seizure onset zone classification using HFO subtype and sharp-spike rates for all patients (N=35) in the designated neuroanatomic regions. Abbreviations (rono: ripples on oscillations, rons: ripples on spikes, frono: fast ripples on oscillations, frons: fast ripples on spikes, AUC: area under the ROC curve). Brackets indicate 95% confidence intervals calculated using bootstrapping (n=1000 surrogates). Please see the figure insets for statistically significant differences in the AUCs between HFO and spike biomarker types and across regions.

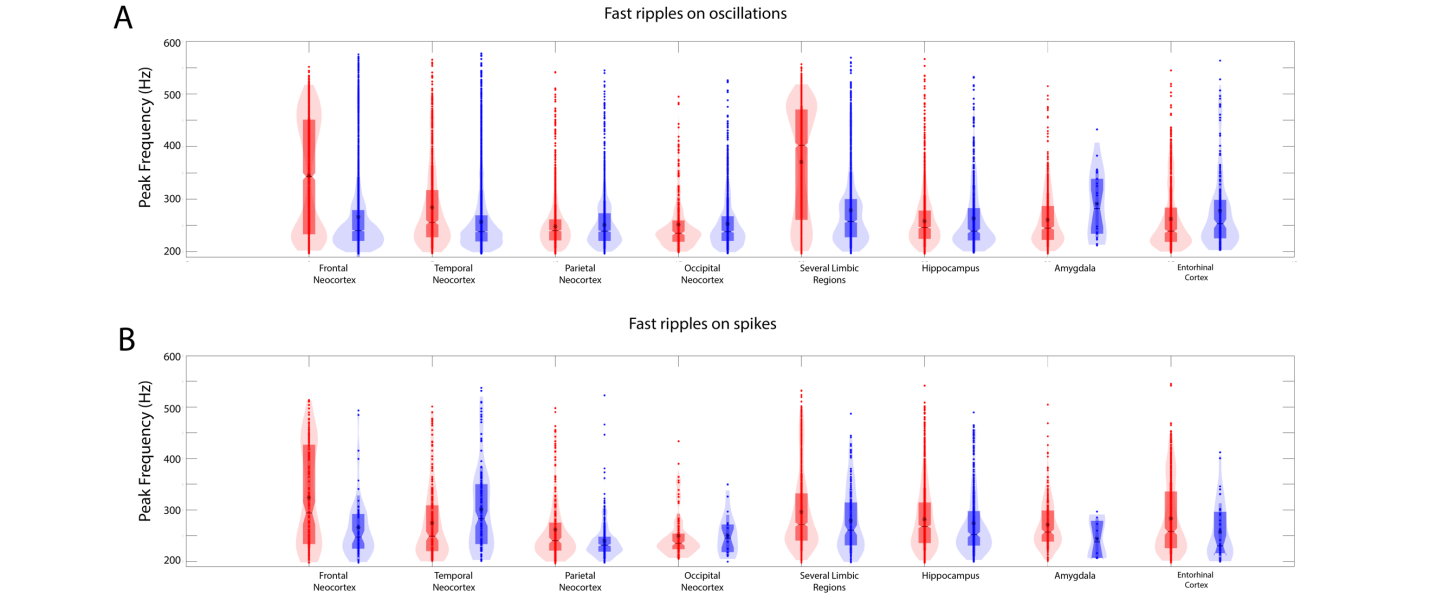

Figure S2: Violin plots of fast ripple on oscillation (A) and fast ripple on spike (B) peak frequency in the seizure onset zone (SOZ, red) and non-SOZ (blue) in all patients for the different designated neuroanatomic locations. Asterisk indicates mean. fRonO and fRonS peak frequency significantly varied by neuroanatomical location (Generalized linear mixed-effects model, GLMM, p<1e-5).

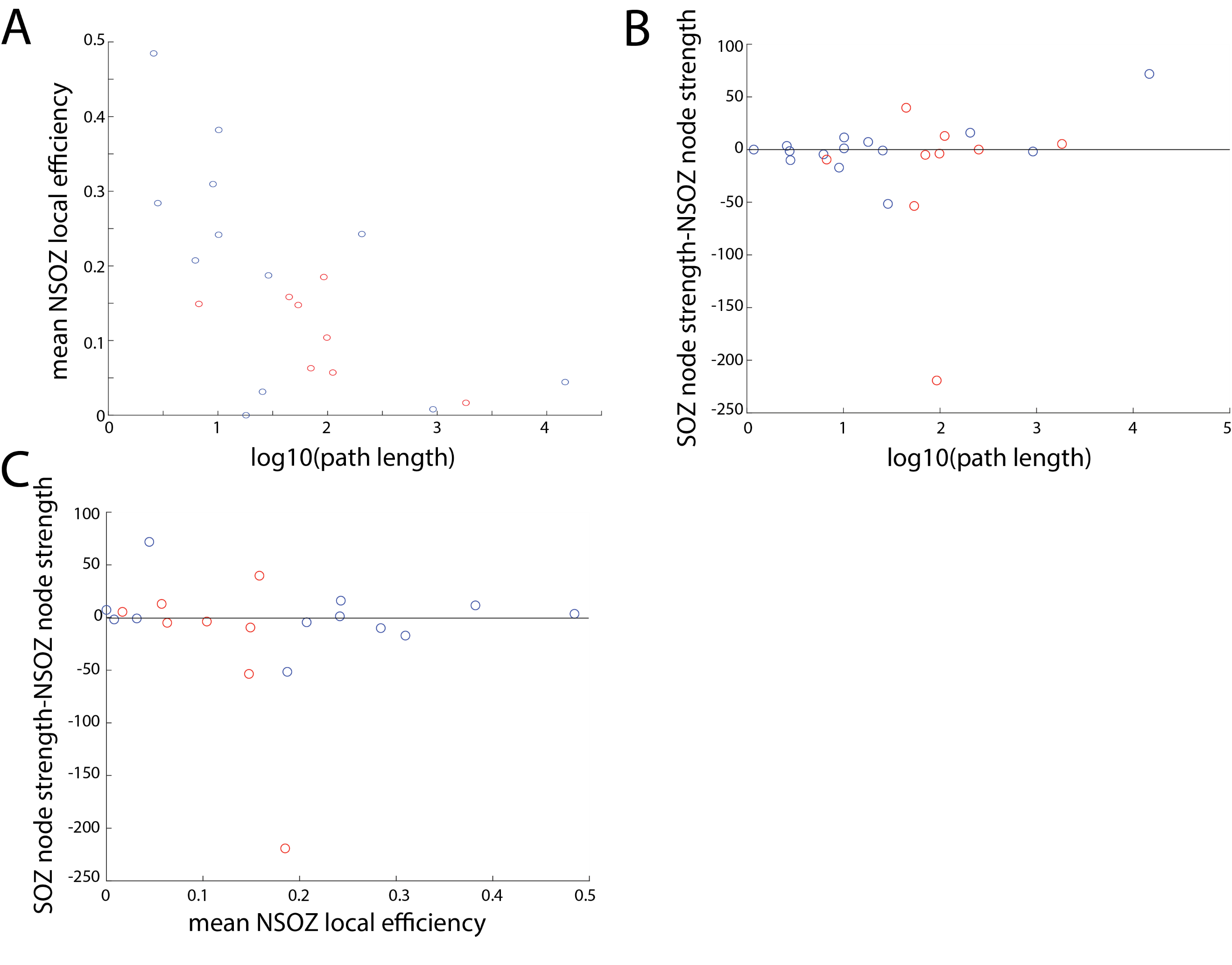

Figure S3: Global metrics of three global graph theoretical measures of the mutual information (MI) networks defined by the fast ripples > 350 Hz. A) Scatter plot of the log transformed path length and the mean non-SOZ (NSOZ) local efficiency for responders (blue) and non-responders (red). B) Scatter plot of the log transformed path length and the difference in summed nodal strength between the SOZ and NSOZ nodes for responders (blue) and non-responders (red). C) Scatter plot of the mean NSOZ local efficiency and the difference in summed nodal strength between the SOZ and NSOZ nodes for responders (blue) and non-responders (red).

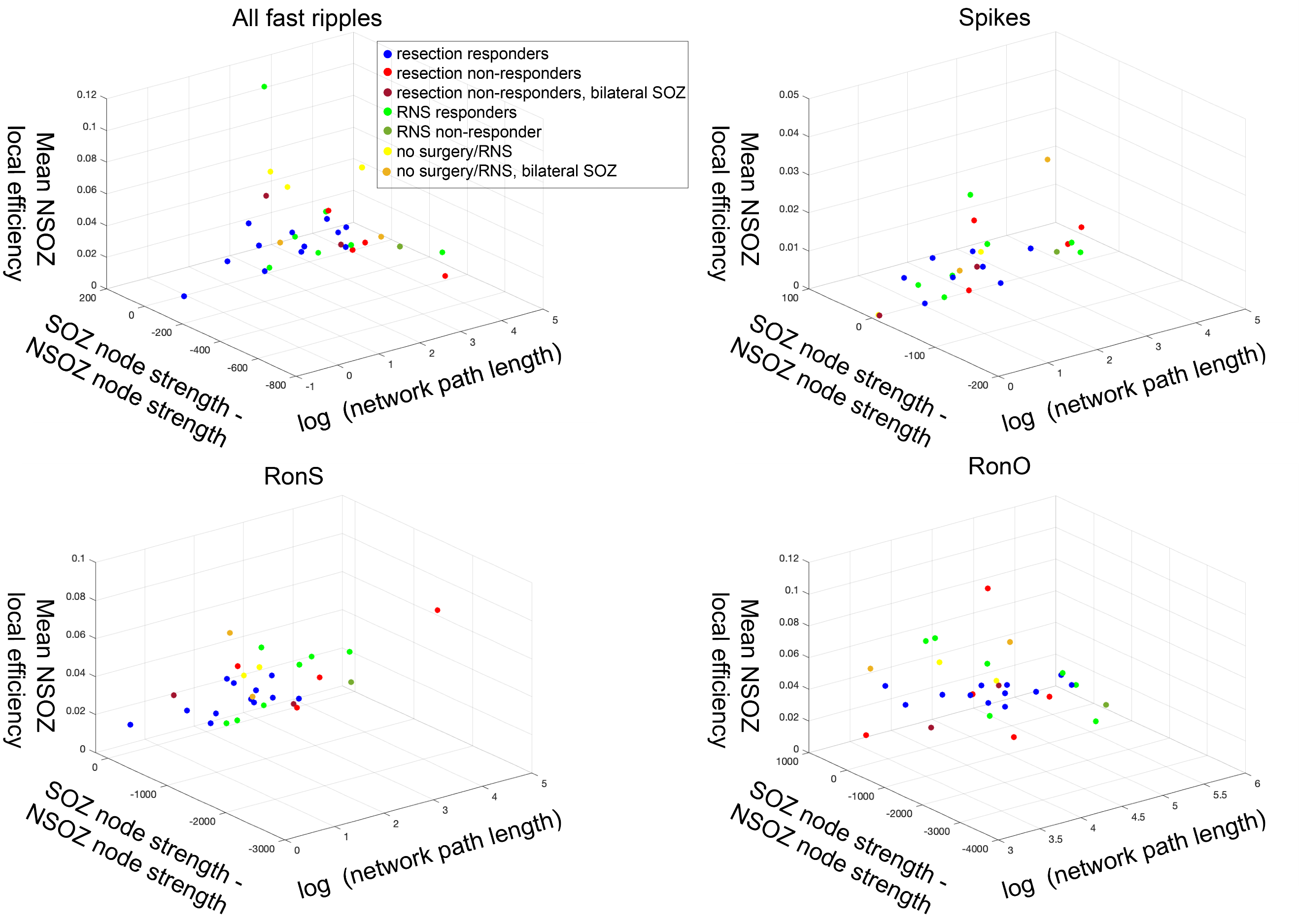

Figure S4: Three-dimensional scatter plot of the three global graph theoretical measures of the mutual information (MI) networks defined by all fast ripples (FR) regardless of frequency content, sharp-spikes, ripples on spikes (RonS), and ripples on oscillations (RonO). Note that the three-dimensional scatter plots of the MI networks defined by the other biomarkers exhibits less clustering of non-responders compared to FR > 350 Hz in Figure 7A.

**Tables**

Table S1: Patient characteristics in the exploratory cohort. Color code: light green corresponds to responders, light red: non-responders and patients not offered resection or RNS, white: excluded from groups due to incomplete follow up. Abbreviations M: male, F: female, L: left, R: right, , N/A: not applicable, ATL: anterior temporal lobectomy, MT(L): mesial temporal lobe, MTS: mesial temporal sclerosis, SMA: supplementary motor area, TBI: traumatic brain injury, LOC: loss of consciousness, RNS: responsive neurostimulator, VNS: vagal nerve stimulator, SUDEP: sudden unexpected death in epilepsy.

| **ID / sex** | **Risk Factor** | **MRI** | **PET (hypo-metabolic)** | **iEEG clinical consensus SOZs** | **Surgery** | **Path.** | **Outcome** |
| --- | --- | --- | --- | --- | --- | --- | --- |
| 1.  2061 M | minor TBI no LOC | Prior frontal lobe resection | N/A | R cingulate gyrus, Medial frontal gyrus, precentral gyrus | RNS medial frontal strip along inter-hemispheric cortex, and strip in dorsolateral frontal lobe behind the resection margin. | N/A | Engel IIB@12 months |
| 2.  4100 M | hypertensive encephalopathy | post L ATL | N/A | L middle temporal gyrus | modified L temporal lobectomy | Gliosis | Engel IVB @48 months |
| 3. 4110 F | encephalitis | Encephalo-malacia | Normal | L inferior frontal gyrus, insula, MT | L temporal lobe and insula resection | Gliosis | SUDEP@6 weeks |
| 4. 4122 M | None | Normal | R temporal | R Inferior temporal gyrus | modified R temporal lobectomy | Gliosis | Engel IA@24 months |
| 5. 4124  F | None | L MTL whitematter hyper-intensity | Normal | R SMA | R frontal lobe resection | cortical dysplasia | Engel IA@24 months |
| 6. 4145 M | None | Normal | Normal | L cingulate gyrus, medial frontal gyrus, middle frontal gyrus, superior frontal gyrus | L frontal lobectomy | cortical dysplasia | Engel IA@40 months |
| 7. 4150 M | significant head injury with LOC | Normal | Normal | R middle frontal gyrus, supplementary motor area , post-central gyrus, pre-central gyrus | R frontoparietal resection, and subpial transection | gliosis | Engel IVB@6 months, lost to follow up. |
| 8. 4163 F | subarachnoid hemorrhage | Encephalo-malacia | L temporal | L pre-central gyrus | L frontal lobe resection | gliosis | EngeI IIIA@42 months |
| 9. 4166 F | meningitis | Encephalo-malacia | L temporal | L MT, uncus, superior temporal gyrus, frontal lesion | L temporal and frontal lobe resection | gliosis | Engel IB@42 months |
| 10. 448 M | infarction | L MTS, encephalo-malacia | L temporal | L fusiform gyrus, middle temporal gyrus, MT, superior temporal gyrus, uncus | L ATL | gliosis | Suicide@10 months |
| 11. 449 M | unknown | Encephalo-malacia | L temporal, parietal | L inferior parietal lobule, post-central gyrus, precuneus, middle temporal gyrus | L parietal lobe resection, RNS post-central gyrus | gliosis | Engel IIA@33 months |
| 12. 451 M | perinatal distress | Normal | L temporal | Bilateral MT, uncus | None | N/A | RNS offered and declined. Lost to follow up. |
| 13. 456 F | None | Normal | R temporal | Bilateral MT, middle temporal gyrus R>L | modified R ATL | gliosis | Engel IVC@48 months |
| 14. 458 M | None | Normal | R temporal | Bilateral MT, uncus. | RNS bilateral entorhinal cortex | N/A | Engel IIIA @ 29 months |
| 15. 463 M | AVM | R occipital AVM | R occipital | Bilateral MT, uncus. | RNS bilateral entorhinal cortex | N/A | Engel IIIA @ 38 months |
| 16. 466 F | None | Normal | L temporal | R fusiform gyrus, superior temporal gyrus, uncus | R ATL | MTS | Engel IB@35 months |
| 17. 468 M | None | L MT FLAIR | R and L temporal | bilateral MT, inferior temporal gyrus, middle temporal gyrus, fusiform gyrus | RNS bilateral entorhinal cortex | N/A | Engel IB @ 24 months |
| 18. 473 F | TBI w/ LOC | L MTS, extra-temporal T2 | L temporal and frontal | L MT, fusiform gyrus, uncus | L MT visualase | N/A | Engel IIIA@18 months |
| 19. 474 F | TBI w /o LOC | vascular malformation | L frontal, temporal, parietal | Left MT, superior temporal gyrus, middle temporal gyrus, inferior temporal gyrus, uncus, frontal lesion | None | N/A | N/A |
| 20. 475 M | head injury LOC unknown | Normal | Normal | bilateral inferior parietal lobule, MT, middle temoral gyrus, inferior temporal gyrus, uncus | VNS | N/A | Engel IVB @ 31 months |
| 21. 477 F | None | periventricular nodular heterotopia, right frontal T2 | R temporal | R MT | ATL | gliosis | Engel IB@31 months |
| 22. 478 F | None | periventricular nodular heterotopia, hypothalamic hamartoma | Normal | bilateral MT, fusiform gyrus, superior temporal gyrus, middle temporal gyrus, inferior temporal gyrus | RNS L entorhinal cortex, L inferior temporal gyrus | N/A | Engel IIB @ 38 months |
| 23. 479 M | TBI w/ LOC | Encephalo-malacia | R temporal | R insula, bilateral middle temporal gyrus, superior temporal gyrus | modified R ATL | gliosis | Engel IVB @ 33 months |
| 24. 480 M | perinatal distress | R hemisphere atrophy, periventricular microgyria | R hemisphere | R fusiform gyrus, insula, MT, middle temporal gyrus, inferior temporal gyrus | R ATL | gliosis | Engel IVC@12 months |
| 25. 481 M | TBI w/o LOC | L MTS | R AND L temporal | L MT, middle temporal gyrus, inferior temporal gyrus, fusiform gyrus, uncus, inferior frontal gyrus, middle frontal gyrus | RNS L entorhinal cortex, L anterior insula | N/A | Engel IIIA @ 33 months |
| 26. IO04 M | None | Normal | L temporal and occipital | L lingual gyrus, middle occipital gyrus | L occipital lobe resection | cortical dysplasia | Engel IA@ 30 months |
| 27. IO05 M | febrile seizures | prior hippocampal sparing temporal lobectomy | N/A | R anterior cingulate, MT, uncus | R ATL | gliosis | Engel IVB@40 months |
| 28. IO06 M | minor head injury no LOC | Normal | N/A | R cingulate gyrus, SMA, post-central gyrus, precuneus, superior parietal lobule | RNS R parietal lobe | N/A | Engel IIIA@ 12 months |
| 29. IO09 M | tuberous sclerosis | L temporal focal cortical dysplasia, nodules, giant cell tumors | L temporal | L MT, fusiform gyrus, superior temporal gyrus, insula, precentral gyrus, inferior frontal gyrus, lesion | None | N/A | Engel IVB@60 months |
| 30. IO10 M | septo-optic dysplastia | R periventricular pachygyria, deficiency of septum pellucidum | N/A | R SMA, frontal lesion, superior parital lobule | right frontal resection subpial transection | gliosis | Engel IVB@35 months |
| 31. IO13 M | None | R parietal lobe resection | R parietal and R occipital | R insula, precuneus, middle occipital gyrus, superior parietal lobule, superior occipital gyrus, superior temporal gyrus, middle temporal gyrus | R parietal | gliosis | Engel IIIA@18 months |
| 32. IO14 M | minor head injury no LOC | Normal | R temporal | L middle temporal gyrus, MT | L ATL | gliosis | Engel IIIA@48 months |
| 33. IO15 M | None | L posterior fossa arachnoid cyst | R temporal | R MT, L cingulate, post. cingulate, mesial frontal, precuneus | R ATL | gliosis | Engel IVB@36 months |
| 34. IO22 M | None | scattered white matter hyperintensities | bilateral temporal hypometabolism | bilateral cingulate gyrus, L middle frontal hyrus, middle frontal gyrus, SMA, pre-central gyrus, post-central gyrus | Anterior corpus callostotomy | N/A | Engel IVB@24 months |
| 35. IO27 F | None | Normal | R temporal | cingulate gyrus, middle temporal gyrus, MT | RNS bilateral mesial-temporal depth eelctrodes | N/A | Engel IVB@18 months |

Table S2: Results of generalized linear mixed-effects models fitting ripple frequency in the different patient cohorts. The random-effect term was the patient, the fixed effects were the SOZ, and the location of the electrode. Brackets indicate 95% confidence interval. Abbreviations n.s. not significant.

| **Response Variable** | **Intercept estimate** | **Intercept p-value** | **SOZ estimate** | **SOZ**  **p-value** | **Location**  **estimate** | **location**  **p-value** | **d.f.** |
| --- | --- | --- | --- | --- | --- | --- | --- |
| All patients RonO freq | 4.680  [4.67 4.69] | <1e-5 | -0.012  [-.013 -.011] | <1e-5 | -0.002  [-.002 -.002] | <1e-5 | 769730 |
| Responders  RonO freq | 4.679  [4.67 4.69] | <1e-5 | -0.006  [-.007 -.005] | <1e-5 | -0.003  [-.004 -.003] | <1e-5 | 431740 |
| Non-Responders RonO freq | 4.681  [4.66 4.70] | <1e-5 | -0.005  [-.007 -.004] | <1e-5 | -0.001  [-.001 -.001] | <1e-5 | 207740 |
| No resction or RNS  RonO freq | 4.721  [4.66 4.77] | <1e-5 | -0.050  [-.052 -.047] | <1e-5 | -0.004  [-.004 .003] | <1e-5 | 84389 |
| All patients RonS freq | 4.685  [4.67 4.70] | <1e-5 | 0.002  [-.001 .006] | n.s. | 7e-4  [-1e-5 2e-3] | n.s. | 55903 |
| Responders  RonS freq | 4.683  [4.67 4.70] | <1e-5 | -0.020  [-.026 -.014] | <1e-5 | 0.005  [.004 .006] | <1e-5 | 21077 |
| Non-Responders RonS freq | 4.677  [4.64 4.71] | <1e-5 | 0.033  [.027 .039] | <1e-5 | -0.003  [-.004 -.002] | <1e-5 | 22172 |
| No resction or RNS  RonS freq | 4.700  [4.64 4.75] | <1e-5 | -0.023  [-.032 .013] | <1e-5 | 0.003  [.001 .004] | .01 | 5918 |

Table S3: Results of generalized linear mixed-effects models fitting HFO subtype power in the different patient cohorts. The random-effect term was the patient, the fixed effects were the SOZ, and the location of the electrode. Brackets indicate 95% confidence interval. Abbreviations n.s. not significant.

| **Response Variable** | **Intercept estimate** | **Intercept p-value** | **SOZ estimate** | **Condition p-value** | **Location**  **estimate** | **location**  **p-value** | **d.f.** |
| --- | --- | --- | --- | --- | --- | --- | --- |
| All patients RonO power | 13.341  [13.2 13.4] | <1e-5 | 0.378  [.370 .387] | <1e-5 | 0.036  [.032 .036] | <1e-5 | 769730 |
| Responders  RonO power | 13.683  [13.5 13.8] | <1e-5 | 0.202  [.189 .215] | <1e-5 | -.0.027  [-.030 -.024] | <1e-5 | 431740 |
| Non-Responders RonO power | 13.109  [12.8 13.4] | <1e-5 | 0.361  [.341 .379] | <1e-5 | 0.084  [.080 .088] | <1e-5 | 207740 |
| No resction or RNS  RonO power | 13.463  [13.1 13.9] | <1e-5 | 0.645  [.602 .688] | <1e-5 | -.010  [-.02 -.001] | 0.03 | 84389 |
| All patients RonS power | 12.173  [11.3 13.0] | <1e-5 | 3.200  [2.71 3.67] | <1e-5 | 0.035  [-.008 .078] | n.s. | 55903 |
| Responders  RonS power | 13.442  [12.3 14.6] | <1e-5 | 2.700  [2.19 3.21] | <1e-5 | -0.171  [-.298 -.044] | 0.008 | 21077 |
| Non-Responders RonS power | 13.051  [12.2 14.0] | <1e-5 | 2.33  [1.86 2.81] | <1e-5 | 0.090  [.060 .120] | <1e-5 | 22172 |
| No resction or RNS  RonS power | 15.573  [15.0 16.2] | <1e-5 | -0.082  [-.270 .103 | n.s. | -0.030  [-.081 .021] | n.s. | 5918 |
| All patients fRonO power | 12.481  [12.4 12.6] | <1e-5 | 0.444  [.399 .488] | <1e-5 | -0.017  [-.026 -.008] | 1e-5 | 41168 |
| Responders  fRonO power | 12.862  [12.7 13.0] | <1e-5 | -.187  [-.318 -.05] | .006 | -0.041  [-.060 -.021] | 1e-5 | 17619 |
| Non-Responders fRonO power | 12.509  [12.3 12.7] | <1e-5 | 0.596  [.544 .648] | <1e-5 | -.006  [-.017 .004] | n.s. | 15283 |
| No resction or RNS  fRonO power | 12.407  [12.2 12.6] | <1e-5 | 0.398  [.331 .464] | <1e-5 | -0.023  [-.043 -.003] | 0.02 | 6022 |
| All patients fRonS power | 12.484  [11.8 13.2] | <1e-5 | 1.964  [1.51 2.41] | <1e-5 | -0.130  [-.208 -.051] | 0.001 | 6302 |
| Responders  fRonS power | 12.782  [11.7 13.9] | <1e-5 | 2.192  [1.61 2.78] | <1e-5 | -0.141  [-.283 .002] | n.s. | 2430 |
| Non-Responders fRonS power | 14.235  [13.6 14.9] | <1e-5 | 0.532  [-.051 1.11] | n.s. | -0.081  [-.163 .002] | n.s. | 2649 |
| No resction or RNS  fRonS power | 15.739  [13.7 17.8] | <1e-5 | -1.537  [-2.6 -.438] | 0.006 | -0.370  [-1.10 .361] | n.s. | 465 |

Table S4: Results of generalized linear mixed-effects models fitting HFO subtype duration in the different patient cohorts. The random-effect term was the patient, the fixed effects were the SOZ, and the location of the electrode. Brackets indicate 95% confidence interval. Abbreviations n.s. not significant.

| **Response Variable** | **Intercept estimate** | **Intercept p-value** | **SOZ estimate** | **SOZ**  **p-value** | **Location**  **estimate** | **Location**  **p-value** | **d.f.** |
| --- | --- | --- | --- | --- | --- | --- | --- |
| All patients RonO duration | -2.992  [-3.0 -2.9] | <1e-5 | -0.022  [-.024 -.019] | <1e-5 | 0.002  [.001 .002] | <1e-5 | 769730 |
| Responders  RonO duration | -2.985  [-3.0 -2.96] | <1e-5 | -0.030  [-.033 -.027] | <1e-5 | 0.002  [.0016 .003] | <1e-5 | 431740 |
| Non-Responders RonO duration | -3.001  [-3.1 -2.9] | <1e-5 | -0.023  [-.027 -.019] | <1e-5 | 0.001  [6e-4 .002] | 1e-5 | 207740 |
| No resction or RNS  RonO duration | -3.021  [-3.1 -2.95] | <1e-5 | 0.002  [-.004 .008] | n.s. | 0.036  [.002 .005] | <1e-5 | 84389 |
| All patients RonS duration | -3.219  [-3.2 3.18] | <1e-5 | -0.007  [-.015 .002] | n.s. | -0.001  [-.003 1e-3] | n.s. | 55903 |
| Responders  RonS duration | -3.149  [-3.2 -3.1] | <1e-5 | 0.005  [-.01 .021] | n.s. | -0.017  [-.02 .013] | n.s. | 21077 |
| Non-Responders RonS duration | -3.267  [-3.3 -3.2] | <1e-5 | -0.039  [-.052 -.025] | <1e-5 | 0.008  [.005 .011] | <1e-5 | 22172 |
| No resction or RNS  RonS duration | -3.261  [-3.3 -3.1] | <1e-5 | 0.014  [-.012 .040] | n.s. | 0.011  [.006 .016] | 1e-4 | 5918 |
| All patients fRonO duration | -4.038  [-4.1 -4.0] | <1e-5 | 0.023  [.01 .036] | 1e-4 | -0.003  [-.005 -6e-4] | 0.014 | 41168 |
| Responders  fRonO duration | -4.117  [-4.2 -4.1] | <1e-5 | 0.100  [0.08 0.12] | <1e-5 | 0.005  [0.001 0.008] | n.s. | 17619 |
| Non-Responders fRonO duration | -4.004  [-4.1 -3.9] | <1e-5 | -0.089  [-.115 -.064] | <1e-5 | -0.006  [-0.01 -.002] | 1e-3 | 15283 |
| No resction or RNS  fRonO duration | -3.948  [-4.0 -3.85] | <1e-5 | 0.058  [.023 .093] | 1e-4 | -0.030  [-.040 -.020] | <1e-5 | 6022 |
| All patients fRonS duration | -4.146  [-4.2 -4.08] | <1e-5 | 0.132  [.093 .170] | <1e-5 | 0.006  [-.002 .013] | n.s. | 6302 |
| Responders  fRonS duration | -4.082  [-4.2 3.99] | <1e-5 | 0.081  [.022 .139] | 0.006 | 3e-4  [-.011 .012] | n.s. | 2430 |
| Non-Responders fRonS duration | -4.133  [-4.2 -4.02] | <1e-5 | 0.155  [0.095 .217] | <1e-5 | 0.001  [-.001 .012] | n.s. | 2649 |
| No resction or RNS  fRonS duration | -4.531  [-4.8 -4.29] | <1e-5 | 0.300  [0.167 0.430] | 1e-5 | 0.046  [0.015 0.077] | 0.004 | 465 |

Table S5: Results of generalized linear mixed-effects models fitting mutual information, local efficiency, and nodal strength. The random-effect term was the patient, the fixed effects were the SOZ, the status of the patient as a responder and non-responder, and the interaction between SOZ and classification as responder and non-responder. Brackets indicate 95% confidence interval.

| **Response Variable** | **Intercept estimate** | **Intercept p-value** | **SOZ estimate** | **SOZ**  **p-value** | **Responder**  **estimate** | **Responder**  **p-value** | **Interaction estimate** | **Interaction p-value** | **d.f.** |
| --- | --- | --- | --- | --- | --- | --- | --- | --- | --- |
| Mutual Information | -0.941  [-1.18 -.7] | <1e-5 | -0.146  [-.32 .02] | 0.093 | 0.231  [-.09 .54] | 0.153 | 0.088  [-.14 .31] | 0.776 | 2496 |
| Local nodal efficiency | -2.22  [-2.7 -1.8] | <1e-5 | -0.20  [-.45 .05] | 0.12 | 0.491  [-.10 1.09] | 0.10 | 0.123  [-.174 .42] | .415 | 218 |
| Nodal  strength | 1.026  [0.5 1.55] | 1e-4 | -0.247  [-.44 -.05] | 0.011 | -0.212  [-.91 .48] | 0.55 | 0.277  [-.08 .63] | 0.12 | 218 |

Table S6: Patient characteristics in the test set cohort. Color code: Red: Patients classified as non-responders by SVM-1 and SVM-2, orange: Patients classified as non-responders by SVM-2 only. Abbreviations: M: male, F: female, L: left, R: right, N/A: not applicable, ATL: anterior temporal lobectomy, MT: mesial temporal, DBS: deep brain stimulation, RNS: Responsive neurostimulator, VNS: vagal nerve stimulator, TBI: traumatic brain injury, LOC: loss of consciousness.

| **ID / sex** | **Risk Factor** | **MRI** | **PET (hypo-metabolic)** | **iEEG clinical consensus SOZs** | **Surgery** | **Path.** | **Outcome** |
| --- | --- | --- | --- | --- | --- | --- | --- |
| 1.  453  M | none | T2 hyperintensity in R temporal pole > L frontal pole. Inferior portion of R temporal pole with blurred gray-white matter border | R temporal | R MT | R temporal ATL | cortical dysplasia IIb | Engel IA@60 months |
| 2.  467 M | none | Blurred gray-white matter border in L inferior parietal lobe | Left inferior parietal lobe | L parietal lobe | L parietal lobe resection and RNS | cortical dysplasia IIb | Engel IIIA@60 months. |
| 3.  470 M | none | L hippocampal atrophy and T2 signal intensity | L temporal | L MT | RNS placed in left MT and entorhinal cortex | N/A | Engel IIB @ 28 months |
| 4. IO01  F | minor TBI | Normal | L temporal | L MT | modified L ATL | Gliosis | Engel IA@24 months |
| 5. IO08 F | other | L temporal lobe atrophy and cortical dysplasia | L temporal | L temporal lobe lesion | modified L temporal lobectomy | cortical dysplasia | Engel IA@40 months |
| 6. IO12  F | none | 1 cm pineal cyst | R lateral temporal | L MT | Modified L temporal lobectomy | Gliosis | Engel IIB@24 months |
| 7. IO17  M | None | Non-specific linear FLAIR signal abnormality in the RT sub-cortical white matter. | R temporal | Left MT, right occipital | Right temporal and occipital resection | Gliosis | Engel IA@36 months |
| 8. IO18  M | Minor TBI | Normal | Normal | Right insula, cuneus, inferior and middle frontal gyrus | R. Frontal lobe | Gliosis | Engel IA @ 24 months |
| 9.  IO19  M | None | Prior R parietal resection | R parietal and occipital hypometabolism | R parietal lobe | R. Parietal lobe resection | Gliosis | Engel IVB @ 36 months |
| 10. IO21  F | None | Prior R. ATL | N/A | Right orbitofrontal cortex. | R. Frontal lobe | hippocampal sclerosis, cortical dysplasia | Engel IVB @ 24 months |
| 11.  IO23 | Significant head injury with LOC | Left temporal T2 hyperintensity with mild enhancement | N/A | Bilateral MT, right lateral temporal | L temporal lobectomy, anterior thalamic DBS | Gliosis | Engel IVB@24 months |
| 12.  IO24 | Significant head injury with LOC | Normal | N/A | R MT, R lateral temporal | None, due to spontaneous improvement after SEEG implant. | N/A | Engel IIB@36 months |
| 13. IO25  F | None | tiny gray matter heterotopia adjacent to the left ventricular atrium, hippocampi appear malrotated | Right temporal hypo-metabolism. | Bilateral MT | VNS | N/A | Engel IVB@24 months |

Table S7: Distance, rate-distance, and information graph theoretical metrics and support vector machine scores for the training set data. Red: non-responder.

|  | SOZ node radius (mm) | Fast ripple node radius (mm) | Fast ripple node rate-distance radius (mm-event/sec) | Fast ripple non-SOZ node rate-distance radius (mm-event/sec) | Characteristic path length | Summed difference in nodal strength of SOZ and non-SOZ nodes | Mean non-SOZ local efficiency | SVM-1 score | SVM-2 score |
| --- | --- | --- | --- | --- | --- | --- | --- | --- | --- |
| '2061' | 58.833 | 61.175 | 11.708 | 11.708 | 6.22 | -4.52 | 0.207 | 0.196 |  |
| '4122' | 45.583 | 52.583 | 2.629 | 2.629 | 0 | 0 | 1 | 0.196 | 0.146 |
| '4124' | 2.591 | 42.353 | 2.641 | 2.641 | 0 | 0 | 1 | 0.196 | 0.153 |
| '4145' | 53.347 | 57.309 | 63.55 | 2.628 | 14701.144 | 71.793 | 0.044 | 0.196 | 0.153 |
| '4163' | 0 | 48.312 | 19.382 | 19.382 | 916.443 | -1.835 | 0.008 | 0.196 | 0.153 |
| '4166' | 22.197 | 78.847 | 11.811 | 11.811 | 25.508 | -0.887 | 0.031 | 0.196 | 0.153 |
| '449' | 50.129 | 57.802 | 1.927 | 0 | 0 | 0 | 1 | 0.196 | 0.153 |
| '458' | 62.12 | 68.348 | 8.543 | 0 | 0 | 0 | 1 | 0.196 |  |
| '463' | 70.427 | 79.163 | 8.923 | 6.001 | 10.167 | 11.517 | 0.382 | 0.196 |  |
| '466' | 43.405 | 90.921 | 4.974 | 6.203 | 2.774 | -1.442 | 1 | 0.196 | 0.173 |
| '468' | 70.043 | 84.872 | 8.521 | 7.671 | 10.142 | 1.151 | 0.242 | 0.488 |  |
| '473' | 7.685 | 65.584 | 9.255 | 8.606 | 1.179 | 0 | 1 | 0.234 | 0.153 |
| '477' | 10.419 | 95.549 | 10.777 | 8.855 | 9.014 | -17.14 | 0.31 | 0.196 | 0.153 |
| '478' | 33.568 | 58.903 | 8.685 | 3.803 | 2.592 | 3.491 | 0.485 | 0.196 |  |
| '481' | 87.536 | 78.022 | 3.997 | 2.43 | 18.035 | 7.225 | 1 | 0.196 |  |
| 'IO004' | 19.755 | 63.574 | 6.701 | 5.947 | 28.897 | -51.556 | 0.187 | 0.224 | 0.153 |
| 'IO006' | 29.381 | 77.221 | 3.293 | 2.481 | 205.483 | 15.995 | 0.243 | 0.196 |  |
| 'IO013' | 38.827 | 82.65 | 5.191 | 5.191 | 2.827 | -10.062 | 0.284 | 0.196 | 0.219 |
| 'IO014' | 21.074 | 84.095 | 5.914 | 5.086 | 0 | 0 | 1 | 0.196 | 0.149 |
| '4110' | 41.008 | 50.583 | 1074.676 | 1074.676 | 1830.626 | 5.357 | 0.017 | 0.680 | 0.929 |
| '479' | 114.832 | 110.979 | 3.329 | 3.329 | 0 | 0 | 1 | 0.746 | 0.930 |
| '480' | 34.589 | 66.999 | 33.488 | 31.62 | 54.003 | -53.541 | 0.148 | 0.463 | 0.294 |
| 'IO015' | 84.451 | 84.967 | 34.493 | 34.616 | 111.686 | 12.917 | 0.057 | 0.792 | 0.937 |
| 'IO027' | 71.194 | 88.53 | 5.827 | 5.827 | 6.701 | -9.484 | 0.149 | 0.721 |  |
| '456' | 19.255 | 69.727 | 9.757 | 6.973 | 251.517 | 0 | 1 | 0.356 | 0.136 |
| 'IO005' | 53.314 | 67.535 | 11.911 | 11.758 | 92.499 | -219.193 | 0.185 | 0.680 | 0.930 |
| '474' | 28.109 | 92.148 | 21.899 | 12.292 | 44.627 | 39.692 | 0.158 | 0.639 | 0.560 |
| '475' | 107.386 | 95.663 | 8.785 | 8.785 | 0 | 0 | 1 | 0.734 | 0.930 |
| 'IO009' | 83.174 | 92.751 | 27.056 | 27.919 | 70.452 | -5.02 | 0.063 | 0.792 | 0.930 |
| 'IO022' | 73.641 | 78.117 | 38.237 | 15.599 | 98.676 | -3.814 | 0.104 | 0.696 | 0.893 |

Table S8: Distance, rate-distance, and information graph theoretical metrics and support vector machine scores for the test set data. Red: non-responder.

|  | SOZ node radius (mm) | Fast ripple node radius (mm) | Fast ripple node rate-distance radius (mm-event/sec) | Fast ripple non-SOZ node rate-distance radius (mm-event/sec) | Characteristic path length | Summed difference in nodal strength of SOZ and non-SOZ nodes | Mean non-SOZ local efficiency | SVM-1 score | SVM-2 score |
| --- | --- | --- | --- | --- | --- | --- | --- | --- | --- |
| 'IO001' | 19.60 | 80.60 | 2.58 | 2.58 | 0.00 | 0.00 | 1.00 | 0.222 | 0.150 |
| 'IO008' | 42.72 | 87.58 | 5.35 | 5.47 | 33.06 | -18.19 | 0.17 | 0.276 | 0.381 |
| 'IO012' | 16.13 | 79.31 | 5.73 | 5.61 | 2.27 | 1.76 | 1.00 | 0.242 | 0.162 |
| 'IO018' | 58.60 | 66.41 | 3.64 | 3.64 | 3.58 | -1.97 | 0.46 | 0.179 | 0.440 |
| 'IO019' | 59.13 | 57.11 | 33.84 | 33.84 | 25.89 | -42.81 | 0.18 | 0.336 | 0.650 |
| 'IO021' | 31.08 | 66.42 | 2.49 | 1.68 | 3.01 | 1.33 | 1.00 | 0.348 | 0.120 |
| '453' | 27.03 | 72.24 | 8.19 | 12.27 | 2.88 | -14.57 | 0.52 | 0.244 | 0.081 |
| '467' | 27.94 | 26.58 | 19.94 | 16.87 | 64.90 | -1.61 | 0.04 | 0.343 | 0.458 |
| 'IO017' | 71.01 | 76.62 | 13.31 | 1.59 | 3.87 | 6.05 | 1.00 | 0.170 | 0.421 |
| 'IO025' | 62.13 | 94.83 | 12.34 | 12.34 | 6.72 | -4.25 | 0.17 | 0.561 | 0.834 |
| '470' | 28.35 | 84.93 | 19.37 | 2.94 | 131.59 | 18.58 | 0.08 | 0.337 | 0.355 |
| 'IO023' | 52.60 | 91.59 | 48.80 | 31.99 | 188.77 | 8.03 | 0.05 | 0.480 | 0.789 |
| 'IO024' | 72.04 | 75.70 | 5.68 | 8.07 | 0.00 | 0.00 | 1.00 | 0.168 | 0.424 |
